# Understanding Inequalities in Mental Health by Family Structure during COVID-19 Lockdowns: Evidence from the UK Household Longitudinal Study

**DOI:** 10.1101/2022.10.27.22281616

**Authors:** Michael J Green, Peter Craig, Evangelia Demou, S Vittal Katikireddi, Alastair H Leyland, Anna Pearce

## Abstract

**Background:** The COVID-19 pandemic increased psychiatric distress and impacts differed by family structure. We aimed to identify mechanisms contributing to these inequalities.

**Methods:** Survey data were from the UK Household Longitudinal Study. Psychiatric distress (GHQ-12) was measured in April 2020 (first UK lockdown; n=10,516), and January 2021 (lockdown re-introduced following eased restrictions; n=6,893). Pre-lockdown family structure comprised partner status and presence of children (<16 years). Mediating mechanisms included: active employment, financial strain, childcare/home-schooling, caring, and loneliness. Monte Carlo g-computation simulations were used to adjust for confounding and estimate total effects and decompositions into: controlled direct effects (effects if the mediator was absent), and portions eliminated (PE; representing differential exposure and vulnerability to the mediator).

**Results:** In January 2021, after adjustment, we estimated increased risk of distress among couples with children compared to couples with no children (RR: 1.48; 95% CI: 1.15-1.82), largely because of childcare/home-schooling (PE RR: 1.32; 95% CI: 1.00-1.64). Single respondents without children also had increased risk of distress compared to couples with no children (RR: 1.55; 95% CI: 1.27-1.83), and the largest PE was for loneliness (RR: 1.16; 95% CI: 1.05-1.27), though financial strain contributed (RR: 1.05; 95% CI: 0.99-1.12). Single parents demonstrated the highest levels of distress, but confounder adjustment suggested uncertain effects with wide confidence intervals. Findings were similar in April 2020 and when stratified by sex.

**Conclusions:** Access to childcare/schooling, financial security and social connection are important mechanisms that need addressing to avoid widening mental health inequalities during public health crises.

**Key Messages:**

- Couples with young children compared to those without, had raised risk of psychiatric distress during UK lockdowns. Effect decompositions suggested this was largely due to a combination of differential exposure and vulnerability to childcare and home-schooling.
- Among those without young children, being single compared to in a couple was associated with raised risk of psychiatric distress during UK lockdowns, with differential exposure to financial strain and loneliness both contributing to this inequality.
- There was not sufficient evidence to indicate that being single with young children increased risk of psychiatric distress relative to couples with young children or singles without.

## Introduction

Psychiatric distress increased globally during the COVID-19 pandemic, especially in countries with higher infection rates and stricter lockdowns^1^. Findings on mental health in the early pandemic^2-10^ differ, with some reporting early growth in anxiety and depression followed by decreases as people adapted, but a pooled analysis of 11 longitudinal population surveys in the UK found increases in psychiatric distress that persisted from March2020-March 2021^11^. Psychiatric distress increased more among: women than men; highly educated than less well-educated people; and adults aged 25-44 than younger and older adults^11^. Unequal impacts may reflect differing exposure to aspects of pandemic management that cause distress, such as: job loss, financial strain, social isolation or caring responsibilities for children or elderly relatives^12^. Inequalities may also reflect differing vulnerability to such mechanisms, or a combination of differential exposure and vulnerability^13^. Understanding how differential impacts develop can help address inequalities.

Family structure may also have been associated with inequalities in exposure and vulnerability to distressing aspects of the pandemic. Lone parents may have experienced greater financial strain than couple parents, for example, because they are more likely to live on a low income^14^, and may have felt more isolated than couples during lockdowns. Working parents also had to balance childcare and home-schooling with work during school closures, and working lone parents had no partner to share these responsibilities.

We explore contributions of five potential mediating mechanisms to inequalities in psychiatric distress by family structure during UK lockdowns: active employment, financial strain, childcare and home-schooling, caring, and loneliness. We compare findings from the 1^st^ UK lockdown in April 2020 (where most existing research is focused), with findings from January 2021 as lockdowns were re-introduced following eased restrictions. Considering reports of women being disproportionally affected by increases in childcare and other caring burdens during the pandemic^15-17^, we examine whether findings differ by gender.

## Methods

### Sample

Understanding Society (the UK Household Longitudinal Study) is a nationally representative longitudinal household panel study, based on a clustered-stratified probability sample of UK households, detailed elsewhere^18^. All adults aged 16+ years in chosen households are invited to participate annually in surveys spanning 24 months. We used pre-pandemic data from 2018-2019, with an individual response rate of 65.4%. Additional online surveys^19^ were conducted during the pandemic and our main analyses use data from surveys in April, Sep and Nov 2020 and Jan 2021 (response rates: 40.3%, 29.2%, 27.3% and 27.2%). We also present descriptive data from surveys in 2017-2018 and in May, June and July 2020.

We defined two analytical samples relating to initial and later UK lockdowns. The analytical sample for April 2020 (1^st^ lockdown) was based on participants aged 16+ from the 2018-2019 pre-pandemic survey who had complete data on all analysis variables from that and the April COVID survey (n=10,516). The January 2021 sample (later lockdown) comprised those participating in the 2018-2019 pre-pandemic survey with complete data for analysis variables from that and from COVID surveys in September and November 2020 and January 2021 (n=6,893; respondent inclusion flowchart in Supplementary Figure S1). Analyses were weighted for survey attrition and non-response.

### Measures

For the April 2020 sample, baseline information was from the 2018-2019 survey, with mediators and outcomes measured in April 2020. For the January 2021 data, baseline information was from either the September 2020 survey (during the period of eased restrictions), or the 2018-2019 survey if not available there. Information for mediating variables was from surveys in November 2020 and January 2021 (see Supplementary Table S2).

### Exposure: Pre-Lockdown Family Structure

Family structure was coded for each individual based on presence of a spouse/partner and children (aged <16 years), as follows: couple with no children; couple with children; single with no children; and single with children.

### Outcome: Psychiatric Distress During Lockdown

Symptoms of anxiety and depression were measured with the 12-item General Health Questionnaire (GHQ)^20^, with scores of 4+ coded as psychiatric distress.

### Mediators

Five mediating pathways reflecting material and psychosocial mechanisms were considered: active employment, financial strain, childcare/home-schooling, caring and loneliness.

Respondents reporting full-time, part-time or self-employment were coded as in active employment, while furloughed respondents were coded as not in active employment.

Financial strain was measured with the question: How well would you say you yourself are managing financially these days? Answers were coded in two categories (living comfortably, doing alright, or just about getting by vs finding it quite or very difficult).

Childcare/home-schooling indicated reporting any time spent on childcare and home-schooling. This was not deterministically related to family structure as some with children aged <16 years did not report spending time on childcare, and some reported spending time on childcare despite not living with a child aged <16 years, for example, they could have been helping someone else or caring for a child aged 16+ years.

Caring indicated looking after or giving special help to someone who was sick, disabled or elderly, either within the same household or outside of the household.

Loneliness was measured with the question: In the last 4 weeks, how often did you feel lonely? Answer categories were: hardly ever or never, some of the time, and often; binary coded to indicate often feeling lonely.

### Confounders

Pre-lockdown confounders included: sex (male vs female), race/ethnicity (white vs non-white ethnic minority), UK country (England, Scotland, Wales or Northern Ireland), age in years (16-34, 35-54, 55+), education (degree-level vs less education), longstanding illness (any vs none); National Statistics Socioeconomic Classification (NS-SEC) codes for occupational class (four categories: professional/managerial; intermediate; routine/unskilled; or long-term non-employed); poverty (household income <60% of the median); smoking status (current vs non-smoker); alcohol consumption measured using the AUDIT-C scale (coded with scores of 8+ indicating moderate-high risk drinking); loneliness (measured as described above); and psychiatric distress (GHQ as above). Additional confounders measured during lockdown included indicators for: changes in family structure from pre-lockdown state; whether respondents had received a letter from the National Health Service advising them to shield/isolate; and whether respondents identified as a keyworkers, i.e. their occupation was considered critical enough for them to continue working in their workplace during the lockdown.

### Analysis

First, we provide descriptive data showing the characteristics of the analytic samples and proportions with high psychiatric distress by family structure over the course of the pandemic. Our main analyses used Monte Carlo g-computation simulations^21^ to decompose inequalities in psychiatric distress by family structure into their mediating mechanisms (details in Supplementary Appendix S3 and Figure S4). Logistic and multinomial models were estimated for analysis variables using observed data on variables assumed to precede them in the causal pathway (Supplementary Figure S5). Models from observed data were then used to simulate hypothetical interventions on family structure and mediating pathways. Simulations were validated by comparing runs with no intervention against the observed data. Comparison of proportions with psychiatric distress across simulation arms with family structure set to different values enabled calculation of risk ratios representing total effects (TE) for each of the following (exposure vs reference) comparisons:

1. Couple with children (exposure) compared against couple with no children (reference); i.e. the effect of children <16 years within couples.
2. Single with no children (exposure) compared against couple with no children (reference); i.e. the effect of being single among those without children <16 years.
3. Single with children (exposure) compared against couple with children (reference); i.e. the effect of being single among those with children <16 years.
4. Single with children (exposure) compared against single with no children (reference); i.e. the effect of children <16 years among those who are single.

Total effects for 1-4 above were decomposed for each mediating mechanism into^22^:

A. A controlled direct effect (CDE) representing the effect of the family structure, in the absence of the mediator.
B. A pure indirect effect (PIE) representing the effect of the family structure, purely through differential exposure to the mediator of interest.
C. A reference interaction (rINT) representing differential vulnerability to the effects of the mediator within the exposure group.
D. A mediated interaction (mINT) representing the combination of differential vulnerability and differential exposure to the mediator.

For ease of presentation, we sum the latter three components (B-D) together into a Portion Eliminated (PE), representing the portion of the effect that would be eliminated in the absence of the mediator, i.e. the portion that is due to differential exposure and/or differential vulnerability to the mediator. Full decompositions are in supplementary material, with relevant insights highlighted in the text. Where controlled direct effects (CDE) are similar to the total effect, this indicates a mediating mechanism does not make a strong contribution. Whereas, if the CDE differs from the total effect and the PE differs from the null, this indicates a contribution to the total effect.

For comparisons 1 and 2, we additionally estimated effects stratified by gender, while this was not possible for comparisons 3 and 4 where the family structure of interest (single with children <16 years) was predominantly female. Additional analyses assessed sensitivity of findings to more conservate confounding assumptions (see Supplementary Figure S6).

## Results

### Descriptive Statistics

Figure 1 shows the prevalence of psychiatric distress by family structure, for two annual pre-pandemic surveys and from surveys at different stages of the pandemic. Mental health was patterned by family structure before the pandemic. Respondents who were single with children had the worst rates of psychiatric distress, followed by those who were single with no children, while those in couples with no children fared best. This patterning was accentuated during the first UK lockdown (covering the April-June 2020 surveys), returned close to previous levels as restrictions eased over summer, and was accentuated again during the further lockdowns.

**Figure 1:**
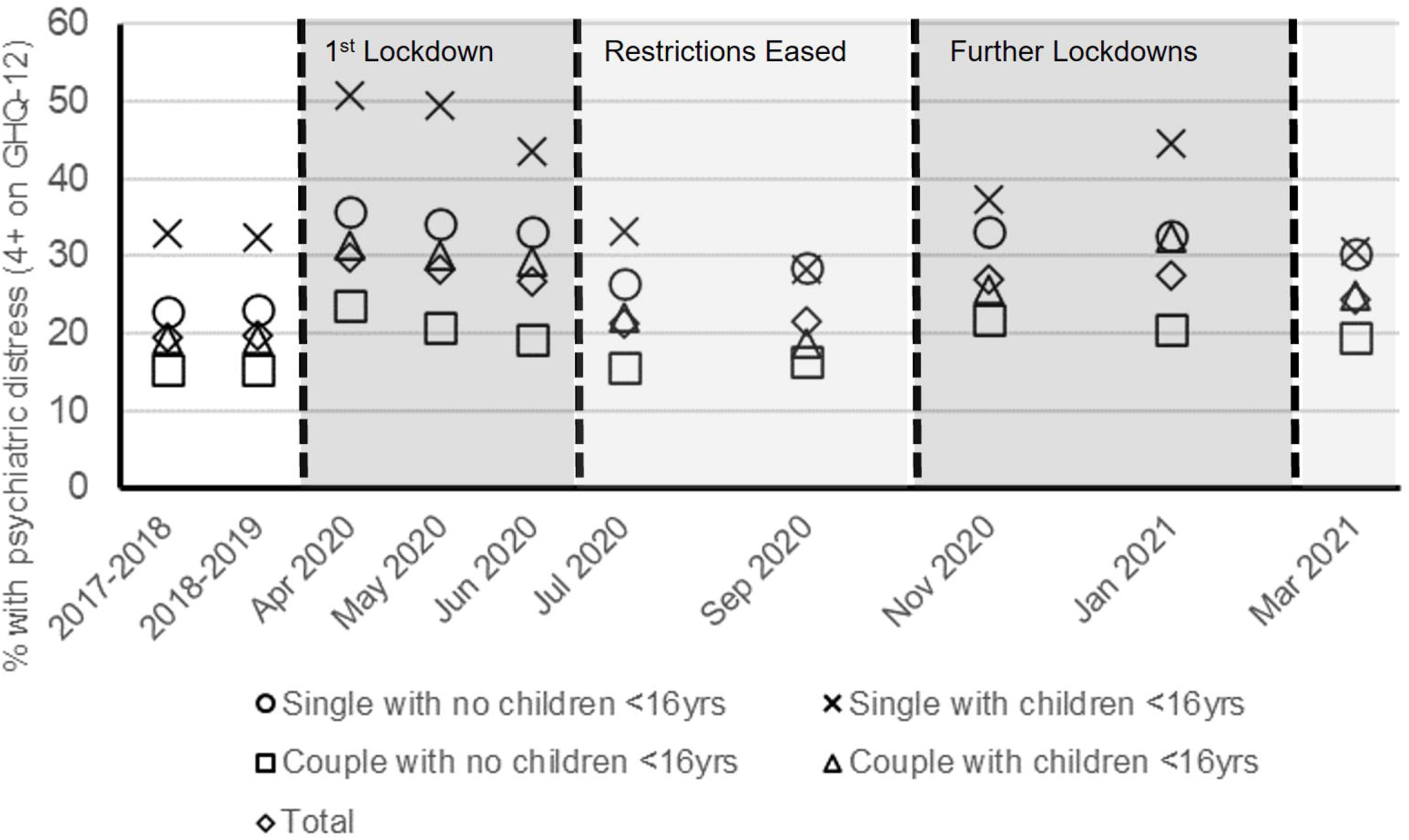
Psychiatric distress by family structure during the COVID-19 pandemic in the UK Household Longitudinal Study.

Table 1 provides descriptive statistics for both samples, overall and by family structure. Single respondents with children were: predominantly female, younger than 55 years of age, more likely to be in household poverty and to be smokers. Single respondents (with or without children) were more likely to often feel lonely, experience psychiatric distress, and report financial difficulties. Respondents with children had higher proportions spending time on childcare and home-schooling, and caring for others in or out of the household.

**Table 1:**
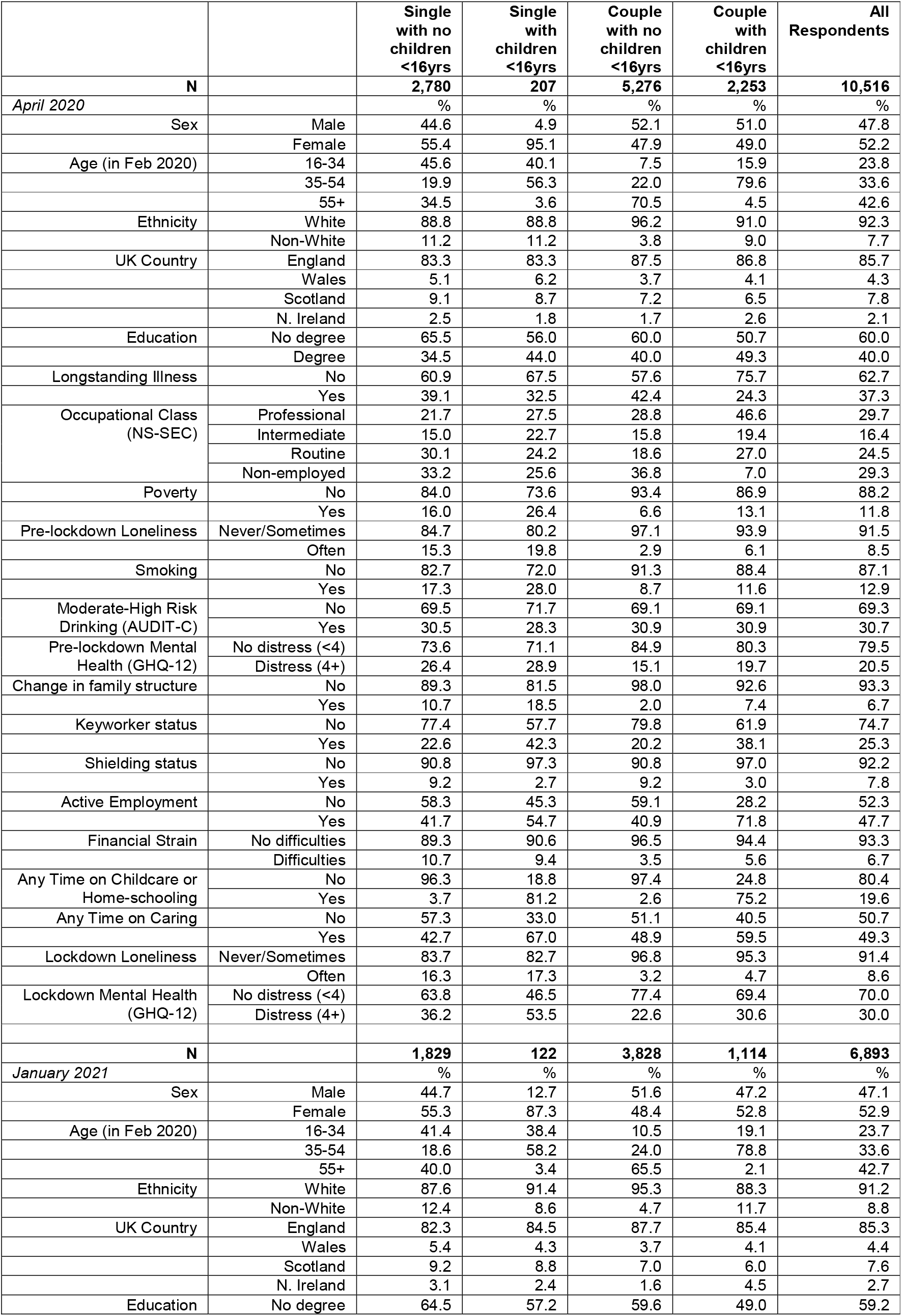
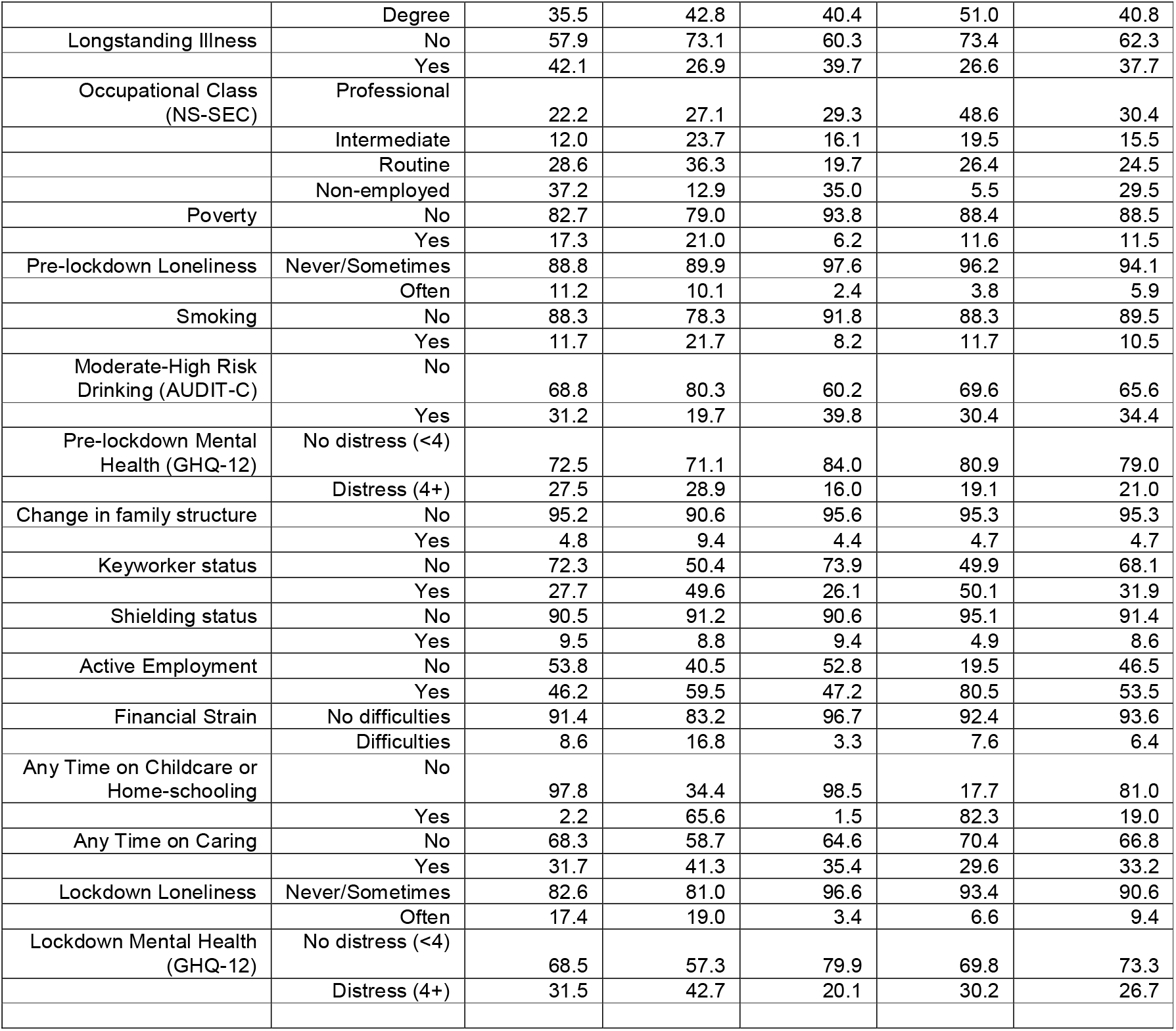
Descriptive statistics by family structure.

Supplementary figures S7-S8 show validation of our simulations comparing observed vs simulated prevalence of all variables (mean and 95% confidence intervals from 1000 simulations), with no manipulation of family structure or mediators. Supplementary figures S9-S16 show further validation, sampling only from within each family structure category. Simulations generally returned proportions similar to those in the observed data.

### Estimated effects of family structure on psychiatric distress and decomposition of effects via mediating pathways

For each comparison, figures display the estimated total effect and decomposition into CDE and PE for each mediating mechanisms in April 2020 and January 2021. Where the PE is closer to the TE and the CDE nearer to the null, the mediating pathway is more important. Supplementary materials detail four-way decompositions, simulated proportions experiencing psychiatric distress in the comparison groups, analyses stratified by gender (where applicable), and sensitivity analyses with more conservative confounding assumptions. Key findings from supplementary material are highlighted below.

#### 1. Couples with children vs. without children

Figure 2 shows the effect estimates (Total, CDE and PE) comparing the risk of psychiatric distress in couples with children against those without. With estimates adjusted for confounding factors, having children was associated with increased risk of psychiatric distress during lockdowns among couples (April 2020 RR: 1.15; 95% CI: 0.98-1.33; Jan 2021 RR: 1.48; 95% CI: 1.15-1.82). In both samples childcare and home-schooling was associated with the largest contributions to the total (April 2020 PE RR: 1.20; 95% CI: 0.99-1.40; January 2021 PE RR: 1.32; 95% CI: 1.00-1.64). The four-way decomposition (Supplementary Table S17) indicated this was mainly due to differential exposure to childcare and home-schooling, but there was evidence of differential vulnerability too, especially in April 2020. In April 2020 couples with children were also less susceptible to remaining in active employment, and that this helped mitigate otherwise larger effects (rINT RR: 0.84; 95% CI: 0.65-1.06 with CDE RR: 1.31; 95% CI: 0.99-1.63), but no similar pattern was apparent in January 2021. When stratifying by gender (Supplementary Tables S18-S19) confidence intervals were wider, but estimates were largely consistent in magnitude with those above, except for females in April 2020 where females in couples with and without children had similarly high proportions with psychiatric distress (36-37%; RR: 1.05; 95% CI: 0.88-1.21). Even here though, there was a clear PE for childcare/home-schooling (RR: 1.30; 95% CI: 1.10-1.49). More conservative control for confounding gave similar findings (Supplementary Table S20).

**Figure 2:**
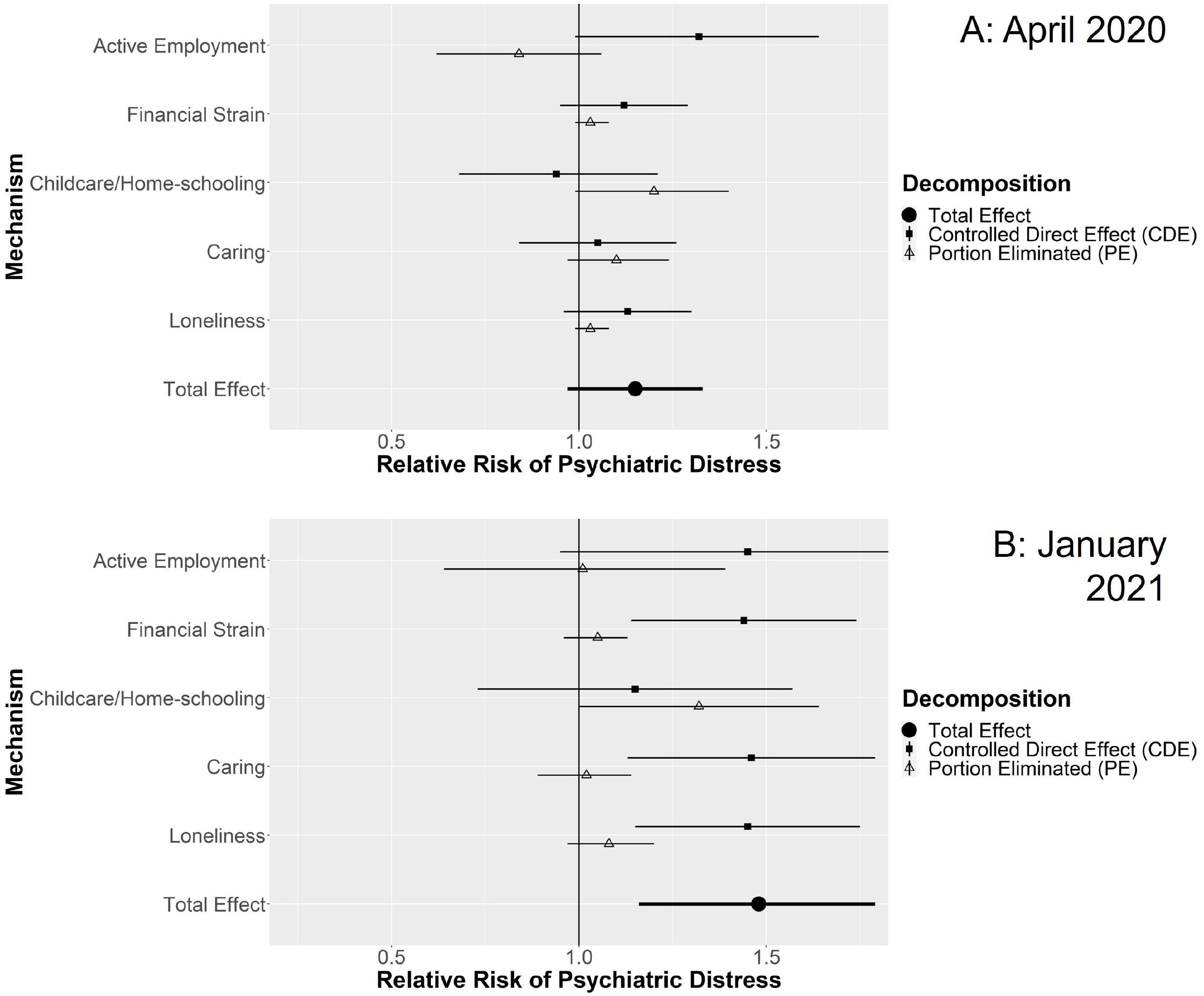
Decompositions for psychiatric distress differences between couples with children vs without children.

#### 2. Singles vs. couples without children

Figure 3 shows estimates for being single compared to being in a couple among those with no children. The confounder adjusted estimates indicated that being single was associated with increased risk of psychiatric distress during lockdowns (April 2020 RR: 1.28; 95% CI: 1.11-1.44; Jan 2021 RR: 1.55; 95% CI: 1.27-1.83). In both April 2020 and January 2021, the largest PEs were related to loneliness (April 2020 RR: 1.18; 95% CI: 1.12-1.24; January 2021 RR: 1.16; 95% CI: 1.05-1.27). Finer decomposition (Supplementary Table S21) indicated this was due to differential exposure to loneliness. There were also minor PEs related to financial strain (April 2020 RR: 1.07; 95% CI: 1.02-1.12; January 2021 RR: 1.05; 95% CI: 0.99-1.12), which in April 2020 were clearly related to differential exposure, while in January 2021 differential exposure may have been combined with differential vulnerability. Results were consistent for both genders (Supplementary Tables S22-S23), except that in January 2021, there was evidence that single compared to couple males experienced differential vulnerability to remaining in active employment (rINT RR: 1.34; 95% CI: 1.00-1.67). Effects were attenuated with more conservative confounding control but remained consistent (Supplementary Table S24).

**Figure 3:**
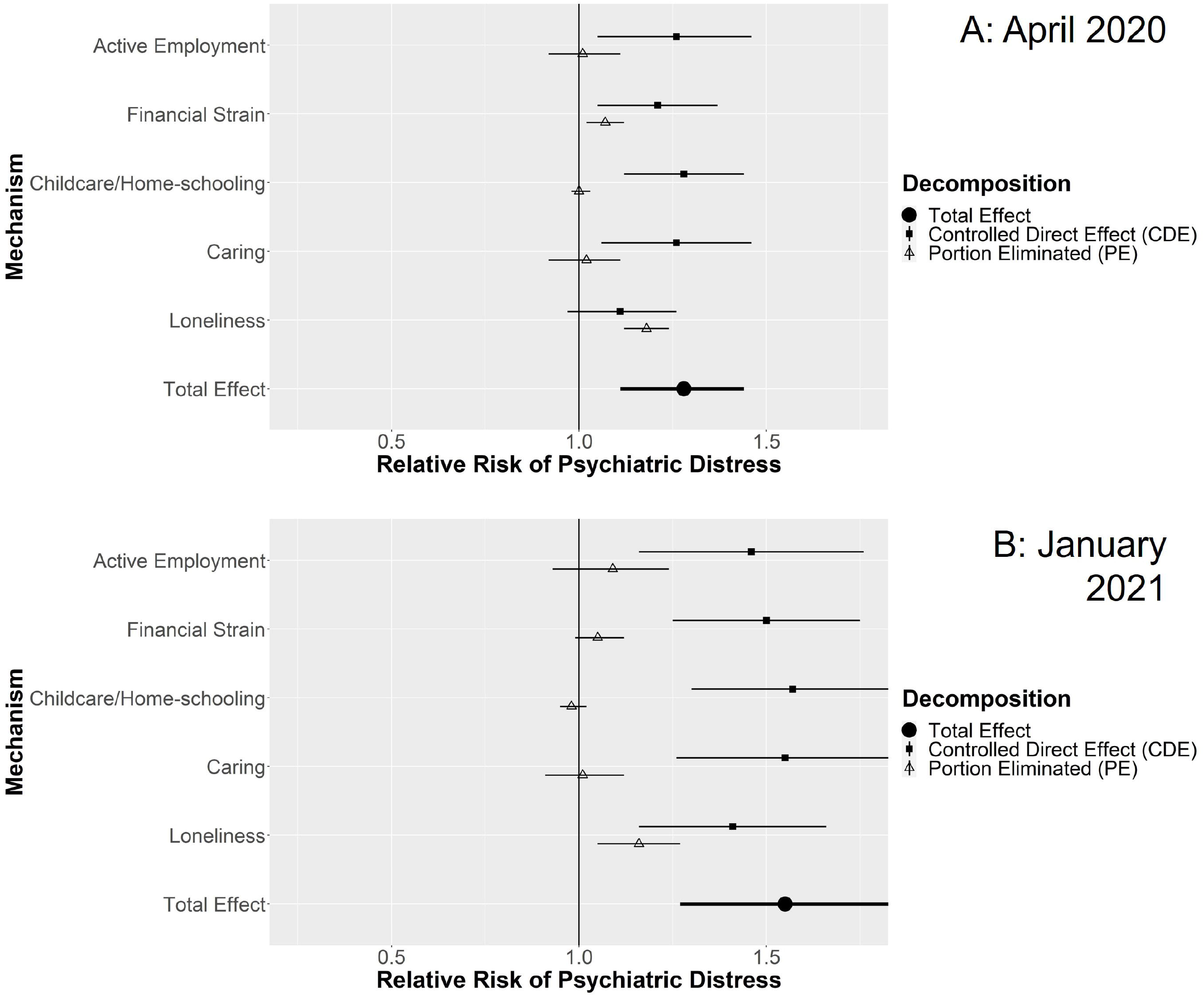
Decompositions for psychiatric distress differences between singles vs. couples without children.

#### 3. Singles vs. couples with children

Estimates in Figure 4 show effects of being single among those with children. An effect could be distinguished from confounding factors in April 2020 (RR: 1.41; 95% CI: 1.03-1.79) but not in January 2021 (RR: 1.22; 95% CI: 0.76-1.69). No mediators clearly contributed to higher rates of distress among single parents, though caring apparently mitigated an otherwise larger effect in January 2021 (RR: 0.79; 95% CI: 0.61-0.98). Detailed decompositions (Supplementary Table S25) suggested single parents were less vulnerable to caring than couple parents. There was evidence of differential exposure to loneliness, but this was mitigated by lower susceptibility such that there was no clear overall PE for loneliness. More conservative confounding control (Supplementary Table S26) still suggested a mitigating effect of lower susceptibility to caring (rINT RR: 0.81; 95% CI: 0.63-0.98), but all other effects were attenuated, including the total effect in April 2020 (RR: 1.26; 95% CI: 0.95-1.57). Thus, differential distress experienced in this group may have been largely due to characteristics established prior to the pandemic (even if these pre-pandemic differences were caused by the difference in family structure).

**Figure 4:**
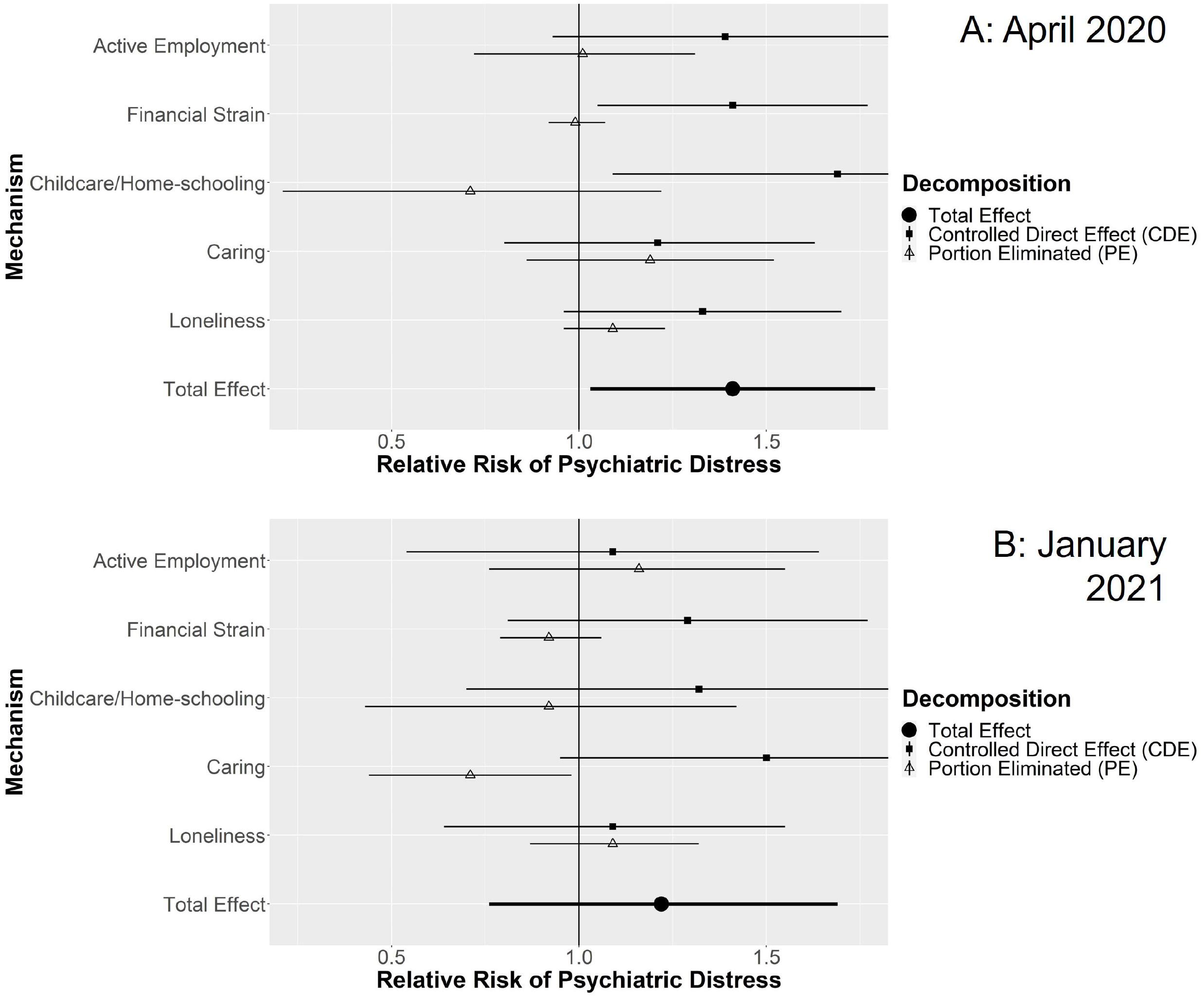
Decompositions for psychiatric distress differences between singles vs. couples with children.

#### 4. Singles with children vs. without children

Estimates in Figure 5 show effects of having children among singles, which could not be confidently distinguished above the effects of confounding factors (April 2020 RR: 1.24; 95% CI: 0.91-1.58; January 2021 RR: 1.10; 95% CI: 0.68-1.52), nor was there clear evidence for contributions from any mediator (Supplementary S27). More conservative confounding control (Supplementary Table S28), also found no clear effects. Thus, differential distress between singles and couples with children appeared largely accounted for by confounding.

**Figure 5:**
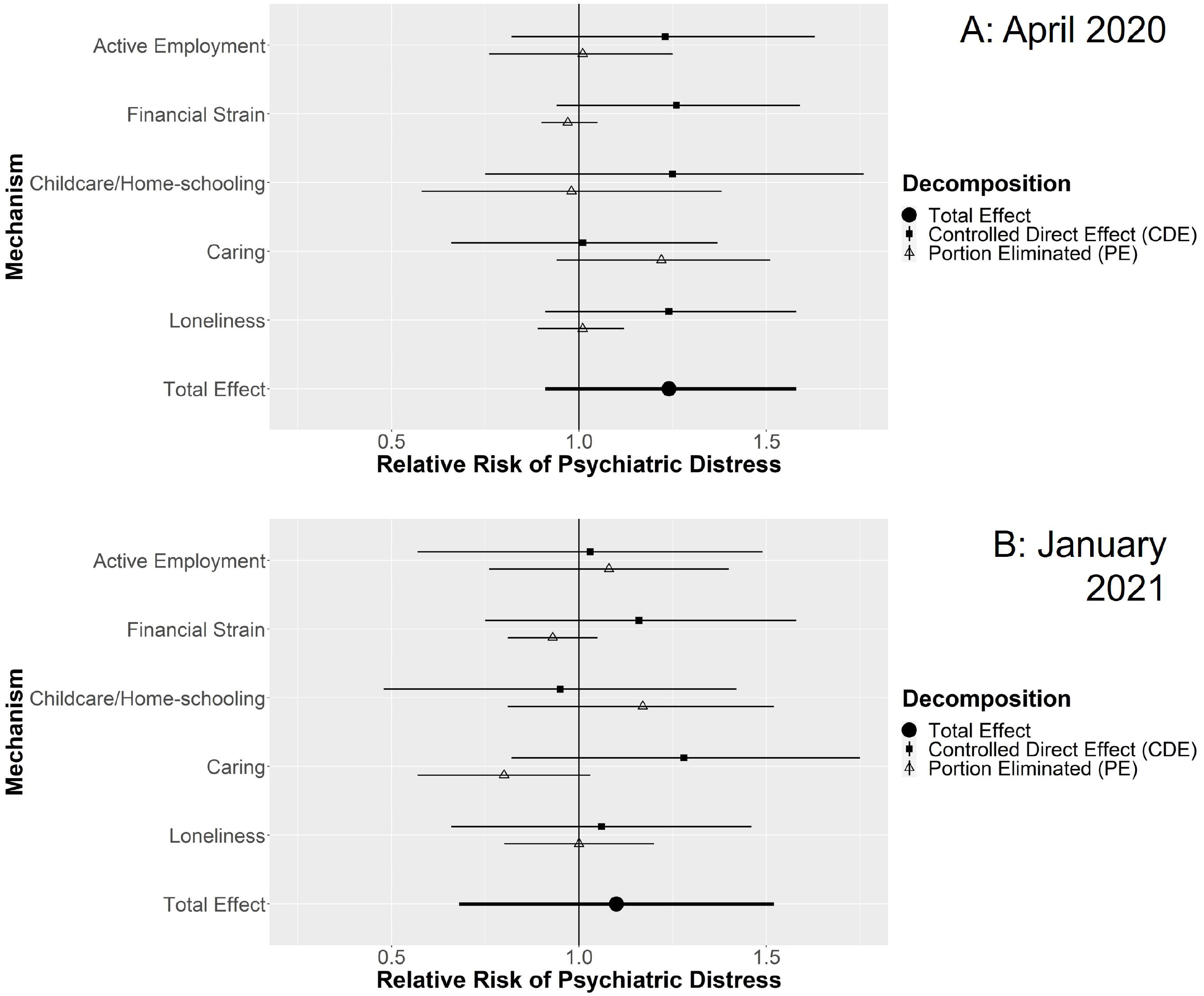
Decompositions for psychiatric distress differences between singles with children vs. without children.

## Discussion

During the UK’s first lockdown in April 2020, and then in January 2021, when lockdowns had been re-introduced after a summer of eased restrictions, we examined differences in psychiatric distress by family structure with representative UK survey data. With a novel simulation approach allowing us to distinguish differential exposure and differential vulnerability to mediating factors, we investigated contributions from active employment, financial difficulties, childcare/home-schooling, other caring, and loneliness. At both time-points, couples with children <16 years had greater risk of psychiatric distress than those without, even after adjusting for confounders. Differential exposure and vulnerability to childcare and home-schooling were the clearest contributing mechanisms. Among respondents who did not have children <16 years, being single rather than in a couple was also associated with increased risk for psychiatric distress at both time-points and after confounder adjustment. Differential exposure to loneliness contributed most clearly to this difference, but differential exposure to financial strain also made a minor contribution. These findings were consistent for both men and women. Single parents (predominantly women) had the highest levels of distress, but effects of single parenthood could not be confidently distinguished above those of confounding factors.

Childcare/home-schooling contributing to differences in mental health between couples with and without children in the home, concur with previous analyses of working parents using the April-May 2020 data^23^, and confirm this was still happening later in the pandemic (January 2021). While facing greater childcare burdens than men during the pandemic may have contributed to higher levels of distress among women^15-17^, we did not find that childcare or caring made greater contributions to the mental health differences associated with having children in the home for women than it did for men. In at least some instances this was apparently because women were experiencing high levels of distress regardless of whether young children were at home. Previous findings also suggest that single parents were especially vulnerable to increased distress during the pandemic^14^, and while we confirmed this descriptively, there was insufficient evidence to support single parenthood as a cause higher distress during the COVID-19 pandemic. This may be due to low statistical power as confidence intervals were wide and relative risk estimates (as well as absolute differences in the estimated proportions with distress) were of similar magnitude to other comparisons. Our estimates were also based on comparing single parents to others with similar background characteristics (i.e. average effects among the exposed), rather than population average effects. Considering that single parents represent a select minority of all families with higher rates of disadvantage, it is feasible that estimates based within this select population could differ from those in the general population, and that risk was raised enough based on other disadvantages that single parenthood had little additional effect. Interestingly, while single parents were more exposed to loneliness than couple parents, they were less vulnerable to it (resulting in no effect overall), suggesting they may have developed adaptive coping strategies for dealing with loneliness.

Causal interpretation of findings rests on assumptions regarding causal direction between variables (as detailed in Appendix S3 and figures S5-S6), perfectly measured confounders, and no unobserved confounders, either of the relationship between family structure and mental health, or of their relationships to mediators. Nevertheless, findings were robust to treating a set of mediator-outcome confounders more conservatively as exposure-outcome confounders. Analyses were weighted for response to COVID-19 surveys, but response rates were relatively low and residual selection bias could remain. Furthermore, estimated contributions of mediating mechanisms to total effects may reflect both differences in the actual strength of causal processes and differences in how well measures reflected any true causal processes.

Data sparsity presented some issues. We could only use a broad age categorisation because of few single parents at older ages. Single parent comparisons exhibited effects sizes of similar magnitude to other comparisons, but confidence intervals were wide so it was hard to distinguish noise from real effects. Additionally, while interactions between family structure and mediators were included, we were not able to examine interactions between mediating mechanisms. For example, childcare and home-schooling might have had more of an effect on distress when coupled with active employment, and other studies have indicated that having to adapt working patterns for such reasons during the pandemic was especially closely linked to distress^24^.

While financial concerns had a minor role in contributing to differences in mental health between singles and couples, neither they nor remaining actively employed, or caring for the sick, elderly or disabled appeared to be strong drivers of inequalities in mental health by family structure. The relative lack of contribution from economic mechanisms may be because the UK job retention scheme (furlough) was helping to ameliorate impacts of economic disruption on mental health^25^. Between the UK’s first lockdown in April 2020 and subsequent lockdowns in late 2020/early 2021 little progress had been made in mitigating key mechanisms leading to family-structure based differences in distress. Demands of childcare and home-schooling appeared to be leading to an excess mental health burden among those with young children in their household, while loneliness was apparently leading to poorer mental health among single people than those in couples. In managing future pandemics or other crises (e.g. involving isolation orders), or even considering on-going moves towards home working, a focus on measures that support financial security, social connections and continued access to childcare, early years’ education and learning, and schooling for young children may be important to avoid exacerbating adult mental health inequalities. Showing who fares worst and why when such resources are curtailed also highlights the importance of these mechanisms for supporting equal population mental health more generally.

## Data Availability

Understanding Society data are available through the UK data service:
Main surveys: https://beta.ukdataservice.ac.uk/datacatalogue/studies/study?id=6614 COVID surveys: https://beta.ukdataservice.ac.uk/datacatalogue/studies/study?id=8644

## Ethics Approval

The University of Essex Ethics Committee approved all data collection. No additional ethical approval was necessary for this study.

## Author Contributions

MJG, PC, ED, SVK, AHL and AP contributed to the research questions and conceptual design of the study. MG conducted all data management, designed and conducted the analysis, and drafted the Methods, Results and Discussion. MJG, PC, ED, and AP reviewed literature and PC drafted the Introduction. MJG, PC, ED, SVK, AHL and AP contributed to revision of the manuscript for critical content.

## Data availability

Understanding Society data are available through the UK data service:

Main surveys: https://beta.ukdataservice.ac.uk/datacatalogue/studies/study?id=6614

COVID surveys: https://beta.ukdataservice.ac.uk/datacatalogue/studies/study?id=8644

## Supplementary data

Supplementary data are available at IJE online.

## Funding

This work was supported by the Medical Research Council [MC_UU_00022/2] and the Scottish Government Chief Scientist Office [SPHSU17]. SVK was additionally supported by a NRS Senior Clinical Fellowship [SCAF/15/02], and AP was additionally supported by a Wellcome Trust University Award [205412/Z/16/Z].

## Conflict of interest

SVK was a member of the UK Scientific Advisory Group on Emergencies (SAGE) subgroup on ethnicity and co-chair of the Scottish Government Expert Reference Group on Ethnicity and COVID-19.

## Notes

### Funding Statement

This study was funded by the Medical Research Council [MC_UU_00022/2] and the Scottish Government Chief Scientist Office [SPHSU17]. SVK was additionally supported by a NRS Senior Clinical Fellowship [SCAF/15/02], and AP was additionally supported by a Wellcome Trust University Award [205412/Z/16/Z].

### Author Declarations

Understanding Society data are available through the UK data service: Main surveys: https://beta.ukdataservice.ac.uk/datacatalogue/studies/study?id=6614 COVID surveys: https://beta.ukdataservice.ac.uk/datacatalogue/studies/study?id=8644

