## Supplementary File 1 for "Understanding Inequalities in Mental Health by Family Structure during COVID-19 Lockdowns: Evidence from the UK Household Longitudinal Study"

Contents:

- Figure S1: Respondent inclusion flow diagram
- Table S2: When measures were taken
- Appendix S3: Description of estimation procedures.
- Figure S4: Analytical Schematic
- Figure S5: Causal assumptions in main analyses
- Figure S6: Causal assumptions in conservative confounding analyses
- Figure S7: Validation Comparison of Observed vs Simulated data: April 2020
- Figure S8: Validation Comparison of Observed vs Simulated data: January 2021
- Figure S9: Validation Comparison of Observed vs Simulated data: April 2020: Single with no children <16 years
- Figure S10: Validation Comparison of Observed vs Simulated data: April 2020: Single with children <16 years
- Figure S11: Validation Comparison of Observed vs Simulated data: April 2020: Couple with no children <16 years
- Figure S12: Validation Comparison of Observed vs Simulated data: April 2020: Couple with children <16 years
- Figure S13: Validation Comparison of Observed vs Simulated data: January 2021: Single with no children <16 years
- Figure S14: Validation Comparison of Observed vs Simulated data: January 2021: Single with children <16 years
- Figure S15: Validation Comparison of Observed vs Simulated data: January 2021: Couple with no children <16 years
- Figure S16: Validation Comparison of Observed vs Simulated data: January 2021: Couple with children <16 years
- Table S17: Decomposition details for couples with children <16 years, compared to couple with no children <16 years
- Table S18: Decomposition details for couples with children <16 years, compared to couples with no children <16 years: Males
- Table S19: Decomposition details for couples with children <16 years, compared to couples with no children <16 years: Females
- Table S20: Decomposition details for couples with children <16 years, compared to couples with no children <16 years: Conservative Confounding
- Table S21: Decomposition details for singles with no children <16 years, compared to couples with no children <16 years
- Table S22: Decomposition details for singles with no children <16 years, compared to couples with no children <16 years: Males
- Table S23: Decomposition details for singles with no children <16 years, compared to couples with no children <16 years: Females
- Table S24: Decomposition details for singles with no children <16 years, compared to couples with no children <16 years: Conservative Confounding
- Table S25: Decomposition details for singles with children <16 years, compared to couples with children <16 years
- Table S26: Decomposition details for singles with children <16 years, compared to couples with children <16 years: Conservative Confounding
- Table S27: Decomposition details for singles with children <16 years, compared to singles with no children <16 years
- Table S28: Decomposition details for singles with children <16 years, compared to singles with no children <16 years: Conservative Confounding

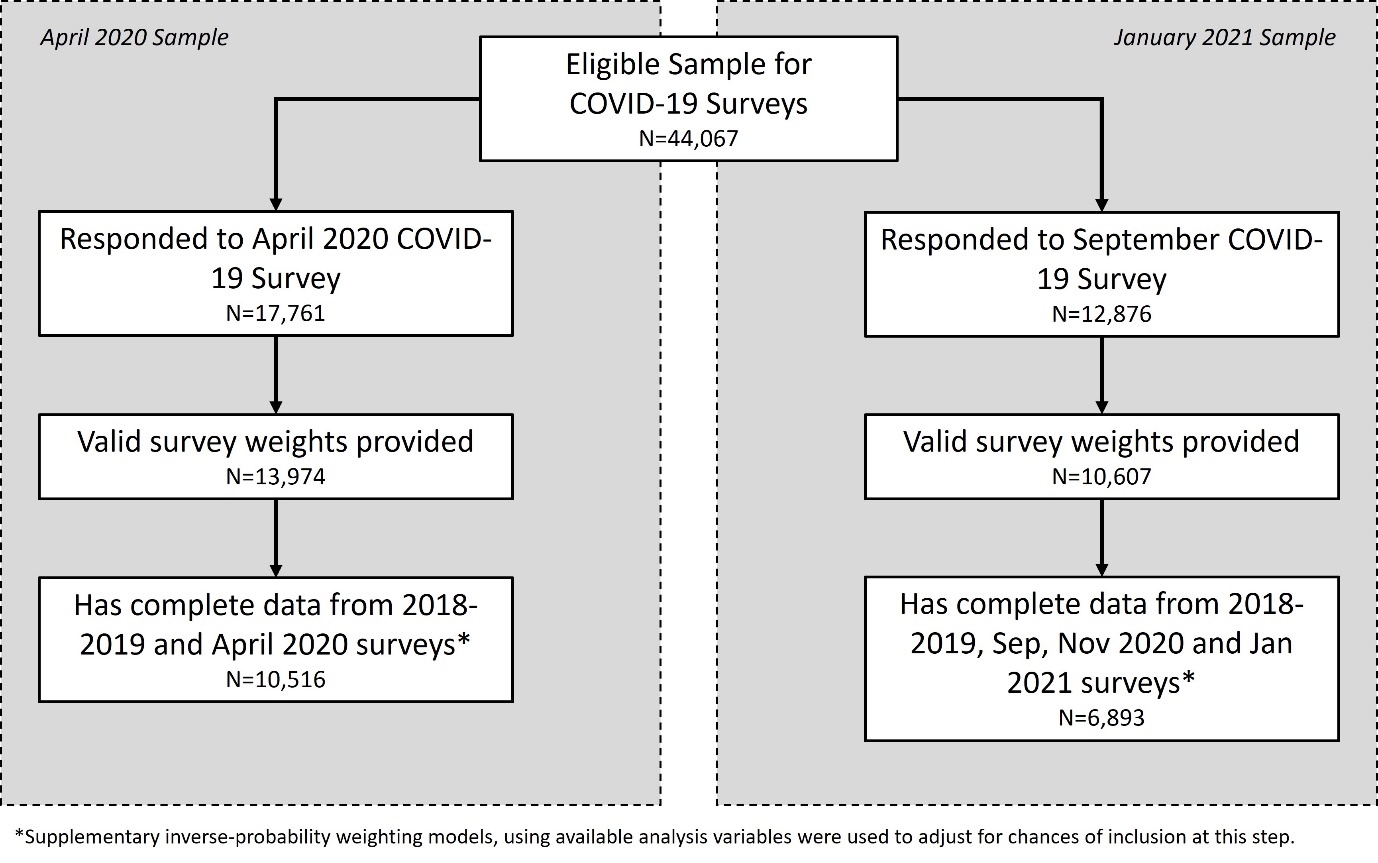

**Figure S1: Participant inclusion flow diagram**

**Table S2: When Measures were taken**

| Measures^a^ | **April 2020 analysis**  N=10,516 | **Jan 2021 analysis**  N=6,893 |
| --- | --- | --- |
| *Pre-Exposure Confounders* |  |  |
| Sex | 2018/2019^b^ | 2018/2019^b^ |
| Age (in Feb 2020) | 2018/2019^b^ | 2018/2019^b^ |
| Ethnicity | 2018/2019^b^ | 2018/2019^b^ |
| UK Country | 2018/2019^b^ | 2018/2019^b^ |
| Education | 2018/2019^b^ | 2018/2019^b^ |
| *Exposure* |  |  |
| Pre-lockdown Family Structure | 2018/2019 | Sep 2020 |
| *Post-Exposure (Pre-Lockdown) Confounders* |  |  |
| Longstanding Illness | 2018/2019^b^ | 2018/2019^b^ |
| Occupational Class (NS-SEC) | 2018/2019^b^ | 2018/2019^b^ |
| Poverty | 2018/2019^b^ | 2018/2019^b^ |
| Pre-lockdown Loneliness | 2018/2019 | Sep 2020 |
| Smoking | 2018/2019 | Sep 2020 |
| Moderate-High Risk Drinking (AUDIT-C) | 2017/2018 | Sep 2020 |
| Pre-lockdown Mental Health (GHQ-12) | 2018/2019 | Sep 2020 |
| *Lockdown Confounders* |  |  |
| Change from pre-lockdown family structure | April 2020 & 2018/2019 | Sep 2020 & Jan 2021 |
| Keyworker status | April 2020 | Jan 2021 |
| Shielding status | April 2020 | April 2020-Jan 2021^c^ |
| *Mediators* |  |  |
| Active Employment | April 2020 | Nov 2020 & Jan 2021^d^ |
| Financial Strain | April 2020 | Nov 2020^e^ |
| Childcare/Home-schooling | April 2020 | Jan 2021 |
| Caring | April 2020 & 2018/2019^f^ | Nov 2020 & Jan 2021^g^ |
| Loneliness | April 2020 | Jan 2021 |
| *Outcome* |  |  |
| Mental Health (GHQ-12) | April 2020 | Jan 2021 |

^a^Assumed causal ordering runs from top to bottom. Although the assumed causal ordering does not exactly match the timing of measurement, especially for the January 2021 analysis, to maintain comparability between the two analyses, we assume that the measures taken are adequate proxies for characteristics that can affect each other in the assumed causal direction, regardless of when measured.

^b^If information was missing, the most recent data from previous annual surveys was used for these characteristics.

^c^Shielding status was based on ever having been advised to shield since the start of the pandemic.

^d^Employment status from November was used first, with status from January used where information from November was missing. There was no question on furlough in November, so furlough information was taken from January.

^e^Measure not available in January 2021.

^f^Information on caring for others outside the household was taken in April 2020 and combined with information on caring for others within the household from the 2018/2019 survey.

^g^Information on caring for others outside the household was taken from the November 2020 survey and combined with information on caring for others within the household from the January 2021 survey.

**Appendix S3: Description of estimation procedures**

Estimation of effects and their decomposition proceeded as follows, with a schematic over-view given in Supplementary Figure S4. R code for the analysis is available in Supplementary File 2. First, we assumed a causal ordering of all variables. This is displayed in Supplementary Table S2 and Supplementary Figure S5, but runs from pre-exposure confounders (sex, age, ethnicity, UK country, education), to the exposure itself (family structure), through post-exposure (but pre-lockdown) confounders (longstanding illness, occupational class, poverty, loneliness, smoking, drinking, mental health), confounders measured during the lockdowns (change in family structure, keyworker status, shielding status), the five mediators of interest (active employment, financial strain, childcare/home-schooling, caring, loneliness), and finally to the outcome (mental health). A key assumption here is the placement of family structure as causally prior to a number of other pre-lockdown characteristics. We therefore additionally present a sensitivity analysis where family structure is placed after these characteristics (i.e. these pre-lockdown characteristics are grouped with the other pre-exposure confounders, see Supplementary Figure S6). We then use the observed data to generate predictive models for all variables that are assumed to occur after the exposure, with each variable predicted by all causally prior variables. These predictive models also accounted for interactions between sex and all other predictors, as well as for interactions between the five mediators and family structure in their effect on mental health. Estimated parameters from these predictive models and their variance-covariance matrix were stored to inform the simulations below.

As family structure had four categories representing the cross-classification of partner status and the presence of children <16 years, we aimed to estimate effects of family structure (and their decompositions) with respect to the following comparisons between an experimental and a reference (control) category:

1. Couple with children (experimental) compared against couple with no children (reference); i.e. the effect of children <16 years within couples.
2. Single with no children (experimental) compared against couple with no children (reference); i.e. the effect of being single among those without children <16 years.
3. Single with children (experimental) compared against couple with children (reference); i.e. the effect of being single among those with children <16 years.
4. Single with children (experimental) compared against single with no children (reference); i.e. the effect of children <16 years among those who are single.

Simulations for each comparison began by re-sampling with replacement from those who had experienced the family structure of interest (probabilities of selection were based on sampling and non-response weights), to create a new sample with the same n and a similar distribution of pre-exposure confounders. This means our estimates are based on the population who actually experienced the family structure of interest, rather than on the general population. That is, we are estimating how that group have been affected by being in that family structure, compared to the counterfactual scenario of that group having been in another reference family structure (i.e. the effect among the treated). For example, since most single parents were young and female, we wanted to estimate effects of single relative to couple parenthood (comparison 3) in a predominantly young, female population.

Following re-sampling from the population of interest, the predictive models described above were used to simulate each post exposure variable in turn, under six distinct experimental manipulation arms (per mediator). These represented:

1. Y0: Family structure set to the reference value (with the mediator taking its natural consequent distribution).
2. Y1: Family structure set to the experimental value (with the mediator taking its natural consequent distribution).
3. Y00: Family structure set to the reference value with the mediator set to 0.
4. Y01: Family structure set to the reference value with the mediator set to 1.
5. Y10: Family structure set to the experimental value with the mediator set to 0.
6. Y11: Family structure set to the experimental value with the mediator set to 1.

Regardless of mediator, the total effect of each family structure is given by comparison of the proportion experiencing psychiatric distress under arm 1 (which we refer to as Y0) and that under arm 2 (referred to as Y1). We express this as a risk ratio (RR; i.e. Y1/Y0).

We further decompose this total effect with respect to each mediator. If we let Y00 denote the simulated proportion of psychiatric distress in arm 3, and similarly, Y01, Y10, and Y11 the same proportion for arms 4-6 respectively, and we let M0 and M1 denote the proportion with the mediating factor present in arms 1 and 2, we decompose into the following four components representing a combination of mediation and moderation^1^:

1. A controlled direct effect (CDE): this represents the effect of the family structure, in the absence of the mediator, i.e. the CDE=Y10-Y00.
2. The pure indirect effect (PIE): this represents the effect of the family structure, purely through differential exposure to the mediator of interest, i.e. the effect of the mediator in the absence of the exposure multiplied by the effect of the exposure on the mediator, or PIE=(M1-M0)*(Y01-Y00).
3. The reference interaction (rINT): which represents differential vulnerability to the effects of the mediator within the exposure group, i.e. the interactive effect of the exposure and the mediator multiplied by the reference prevalence of the mediator, or rINT=M0*(Y11-Y10-Y01+Y00).
4. The mediated interaction (mINT): which represents the combination of differential vulnerability and differential exposure to the mediator, i.e. the interactive effect of the exposure and the mediator multiplied by the effect of the exposure on the mediator, or mINT=(M1-M0)*(Y11-Y10-Y01+Y00).

Following VanderWeele^1^, we express each as a risk ratio, scaled (by Y00/Y0) so that they sum to the total relative risk (i.e. Y1/Y0). For simplicity of presentation, we group the latter three components together into a Portion Eliminated (PE=PIE+rINT+mINT; i.e. the portion of the effect of family structure that would be eliminated in the absence of the mediator) and provide the full four-way decomposition as supplementary material, highlighting any insights from the full decomposition in the text.

Decompositions were performed separately for each mediator, allowing for mediators to influence each other in the assumed causal ordering described above. While causal interpretation of this decomposition normally requires the cross-world independence assumption, i.e. that there be no confounders of the mediator-outcome relationship that are affected by the exposure (as well as usual assumptions regarding no unmeasured confounding of the exposure, mediator and outcome relationships), we avoid the need for this requirement by integrating information on exposure and mediator effects (from arms 3-6) with information on the effect of the exposure on the mediator (from the separate experiments in arms 1-2).

Each simulation run permutated all the predictive model parameters using a random draw from their respective variance-covariance matrices. We report mean values from 1000 simulations with confidence intervals defined by the standard deviation of the simulation results. All effects were estimated separately for both the April 2020 and January 2021 analysis samples.

1.	VanderWeele TJ. A unification of mediation and interaction: a four-way decomposition. *Epidemiology* 2014; **25**: 749-61.

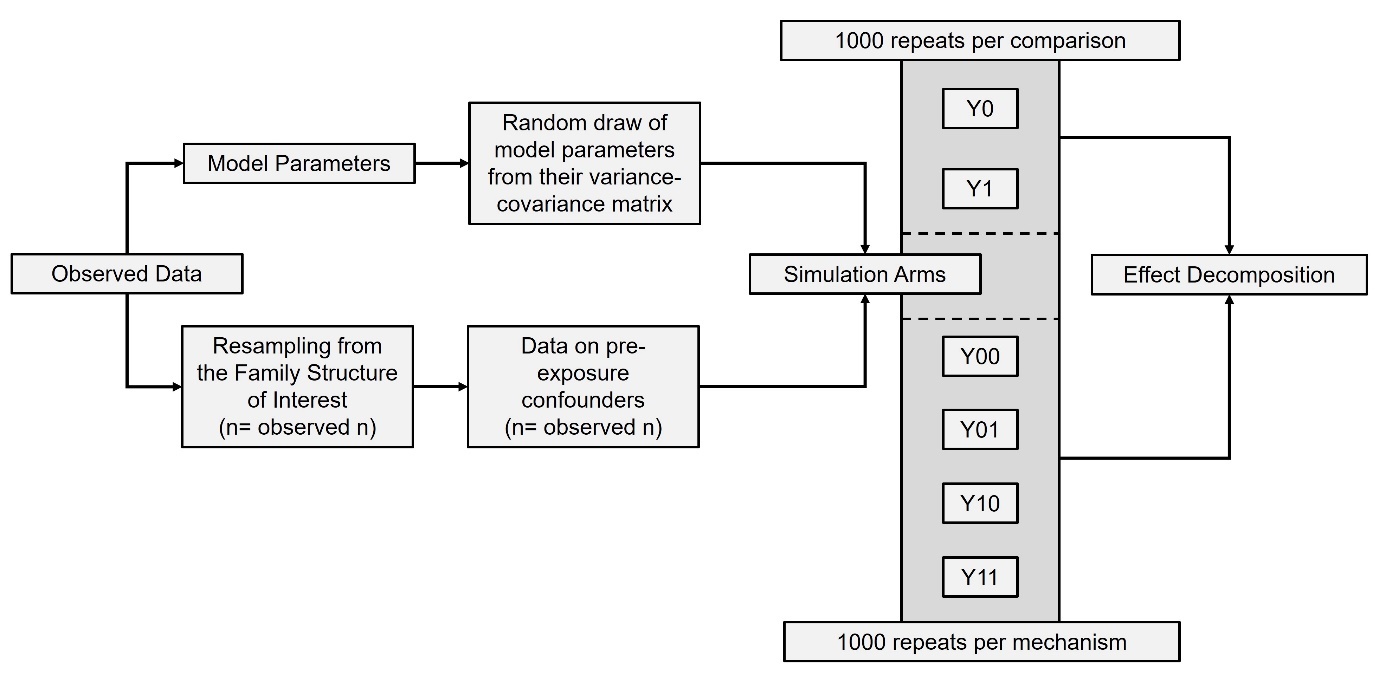

**Figure S4: Analytical Schematic**

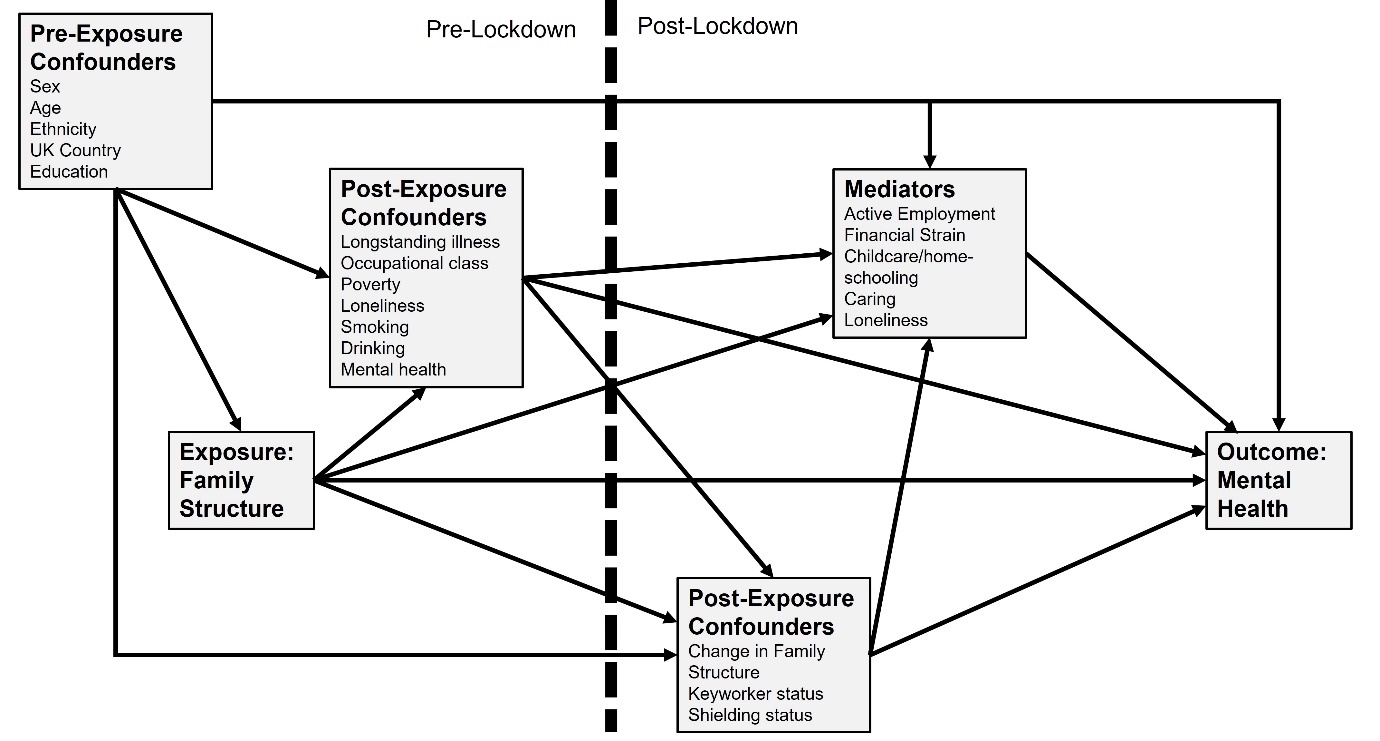

**Figure S5: Causal assumptions in main analyses**

Assumed causal ordering is from top to bottom within each box.

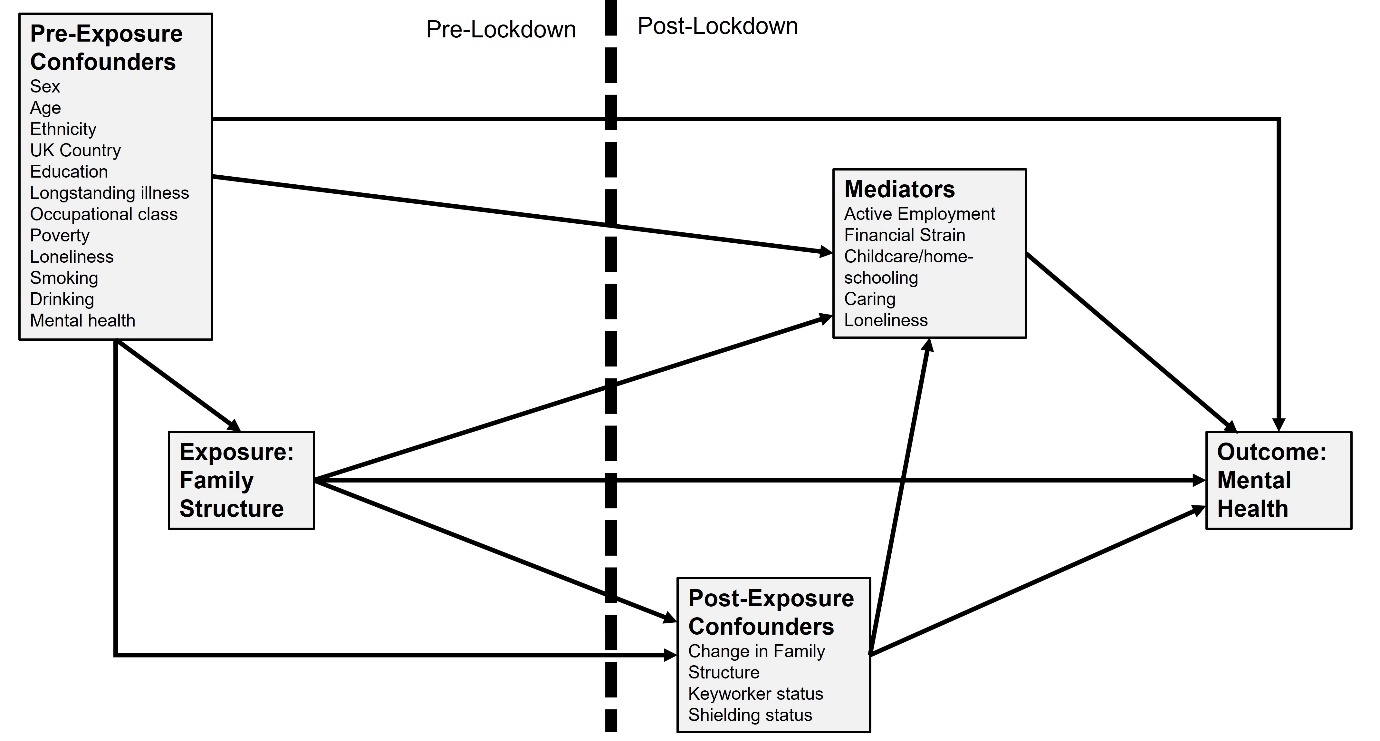

**Figure S6: Causal assumptions in conservative confounding analyses**

Assumed causal ordering is from top to bottom within each box.

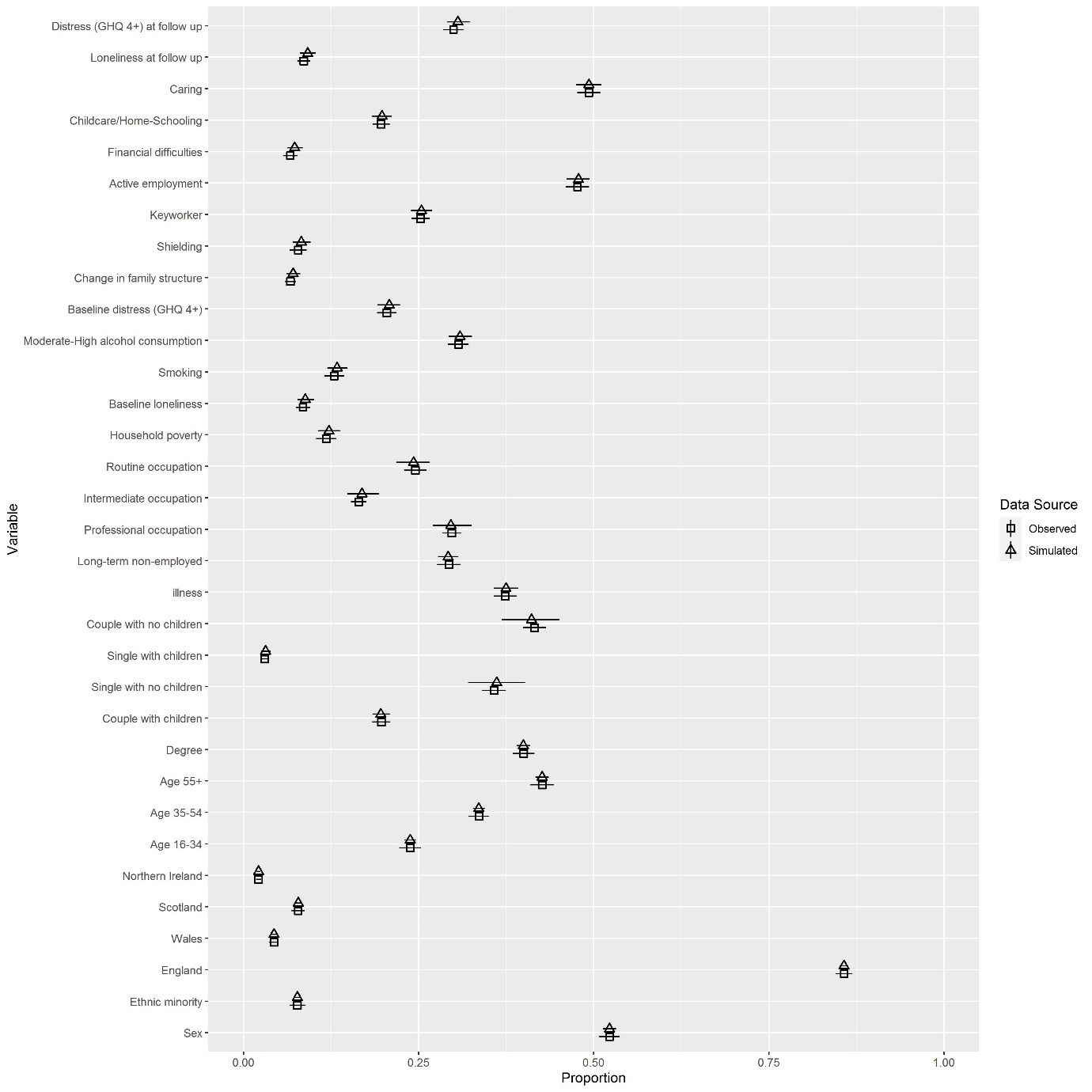

**Figure S7: Validation Comparison of Observed vs Simulated data: April 2020**

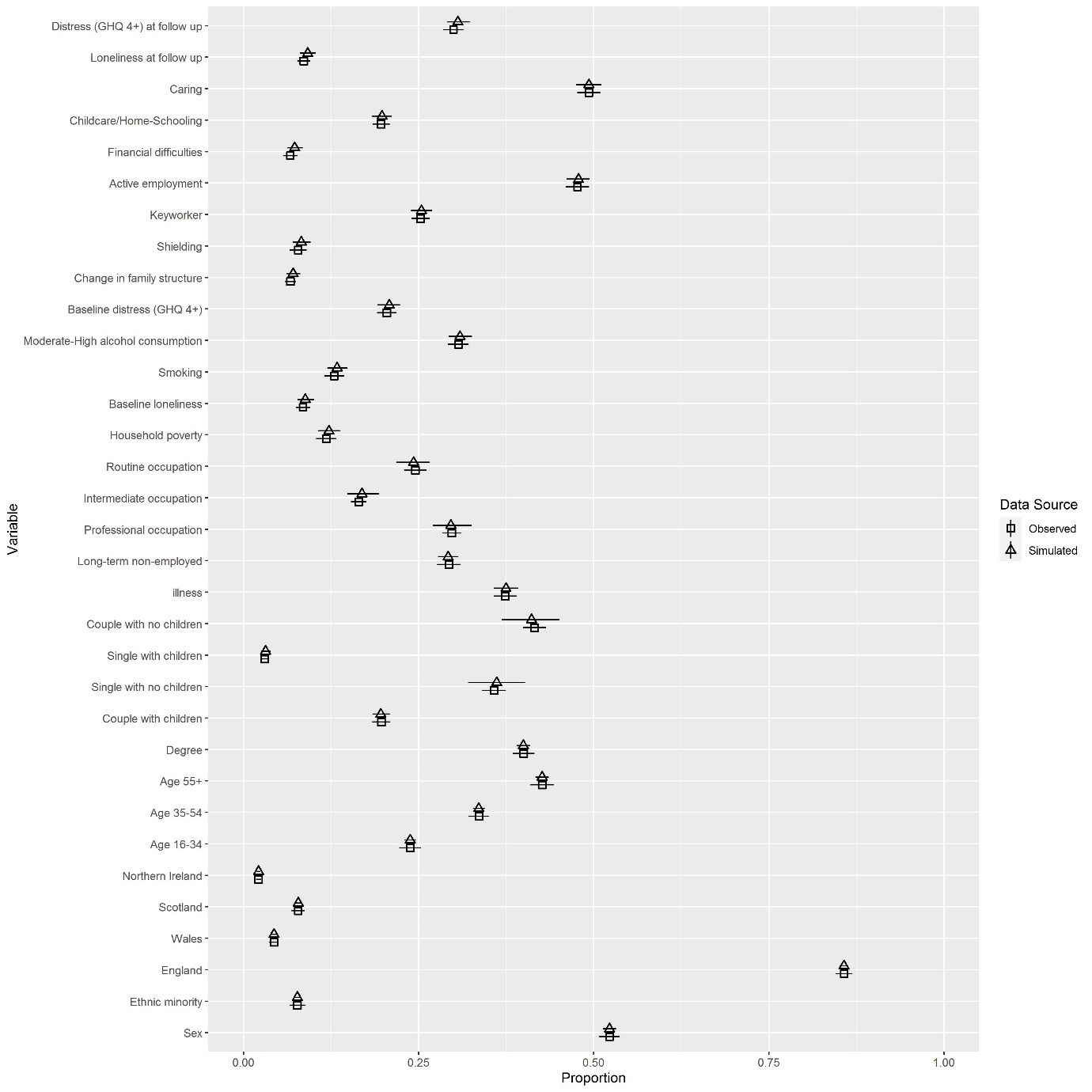

**Figure S8: Validation Comparison of Observed vs Simulated data: January 2021**

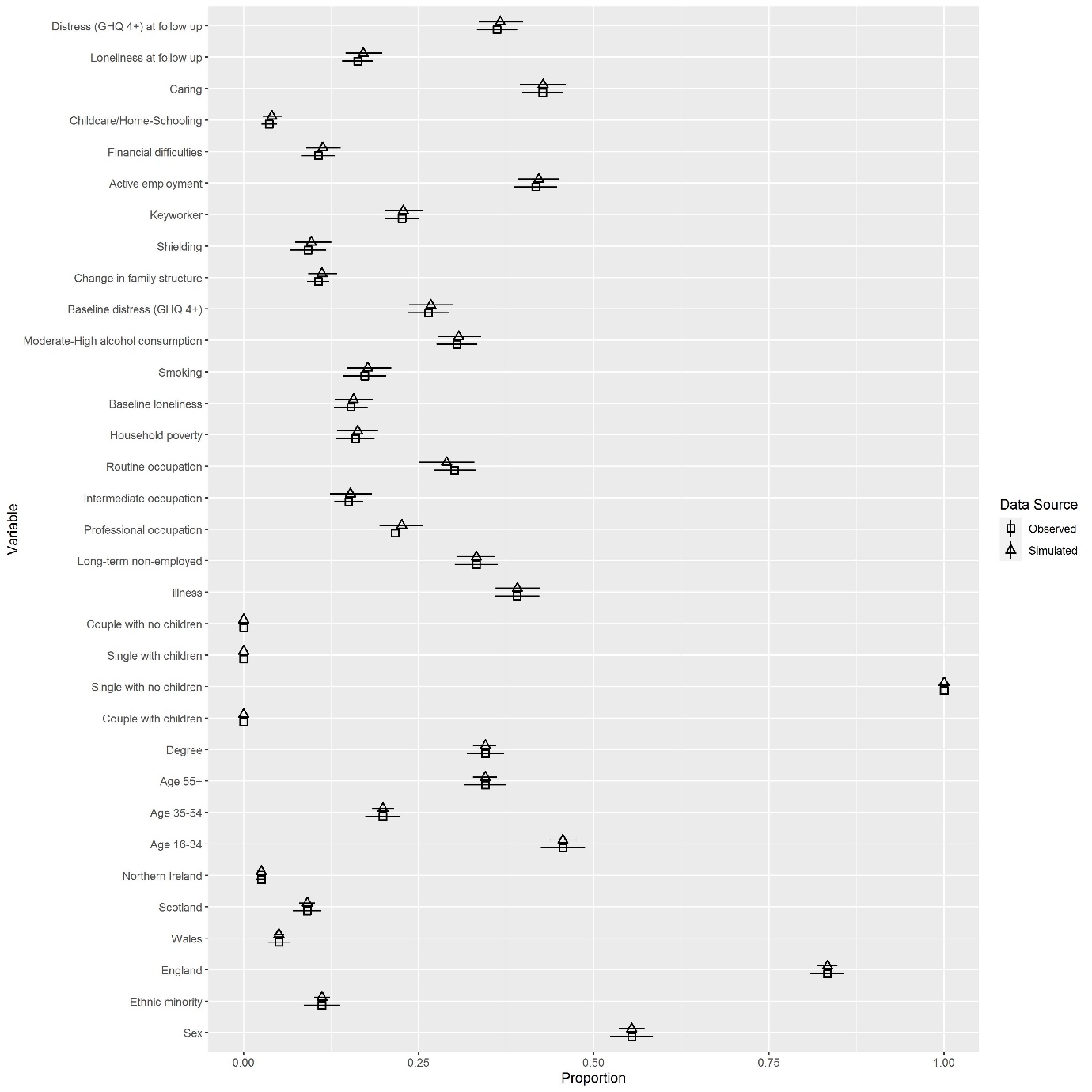

**Figure S9: Validation Comparison of Observed vs Simulated data: April 2020: Single with no children <16 years**

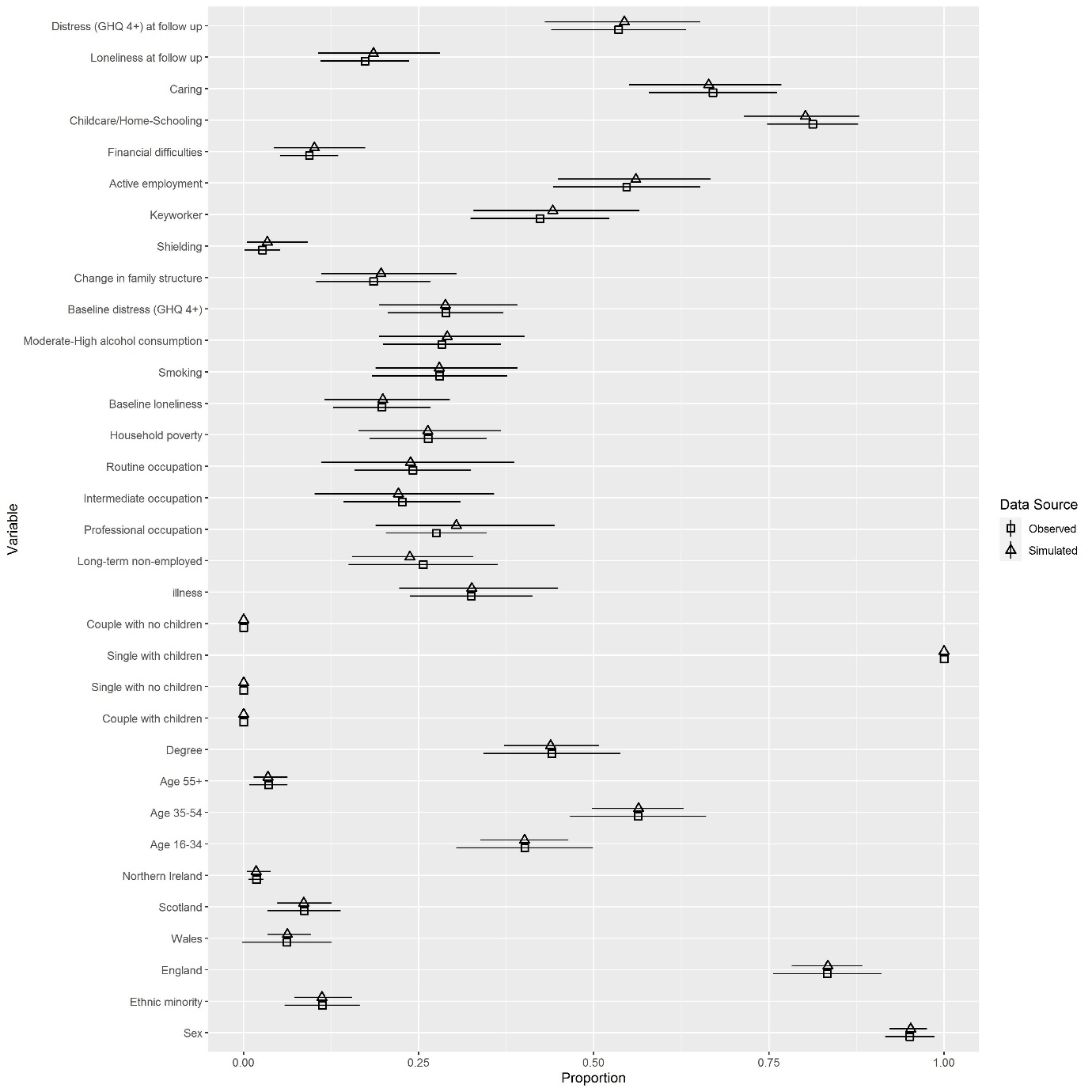

**Figure S10: Validation Comparison of Observed vs Simulated data: April 2020: Single with children <16 years**

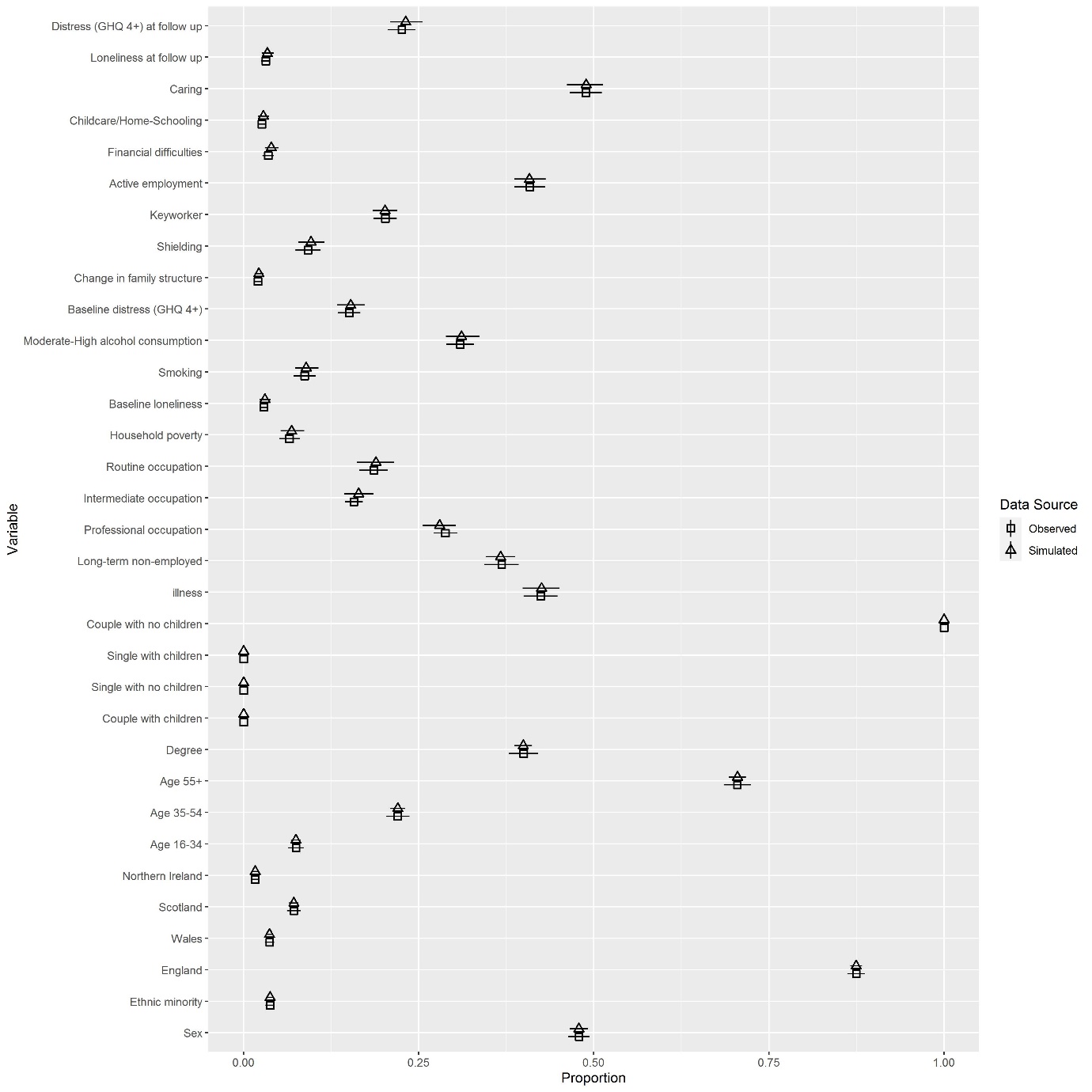

**Figure S11: Validation Comparison of Observed vs Simulated data: April 2020: Couple with no children <16 years**

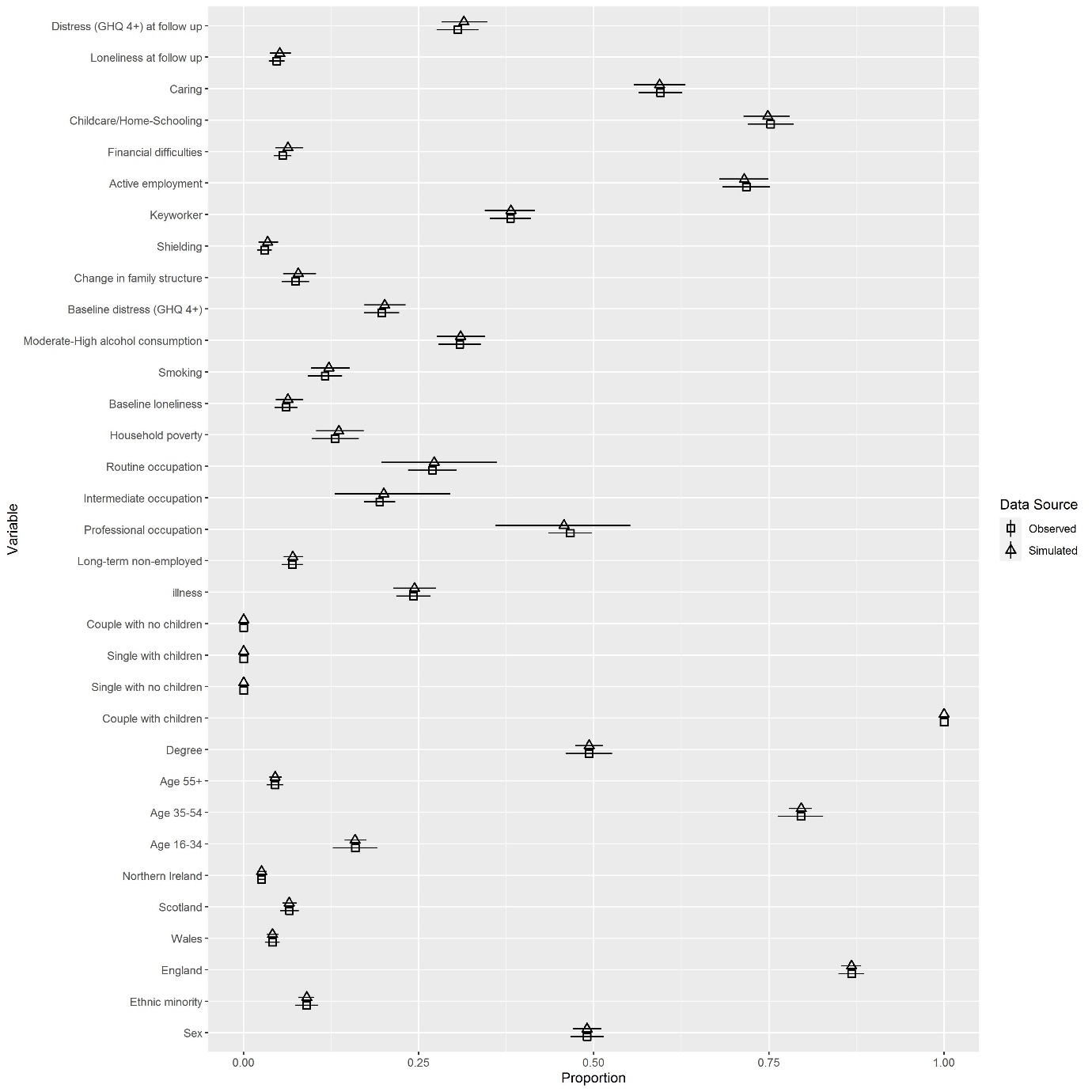

**Figure S12: Validation Comparison of Observed vs Simulated data: April 2020: Couple with children <16 years**

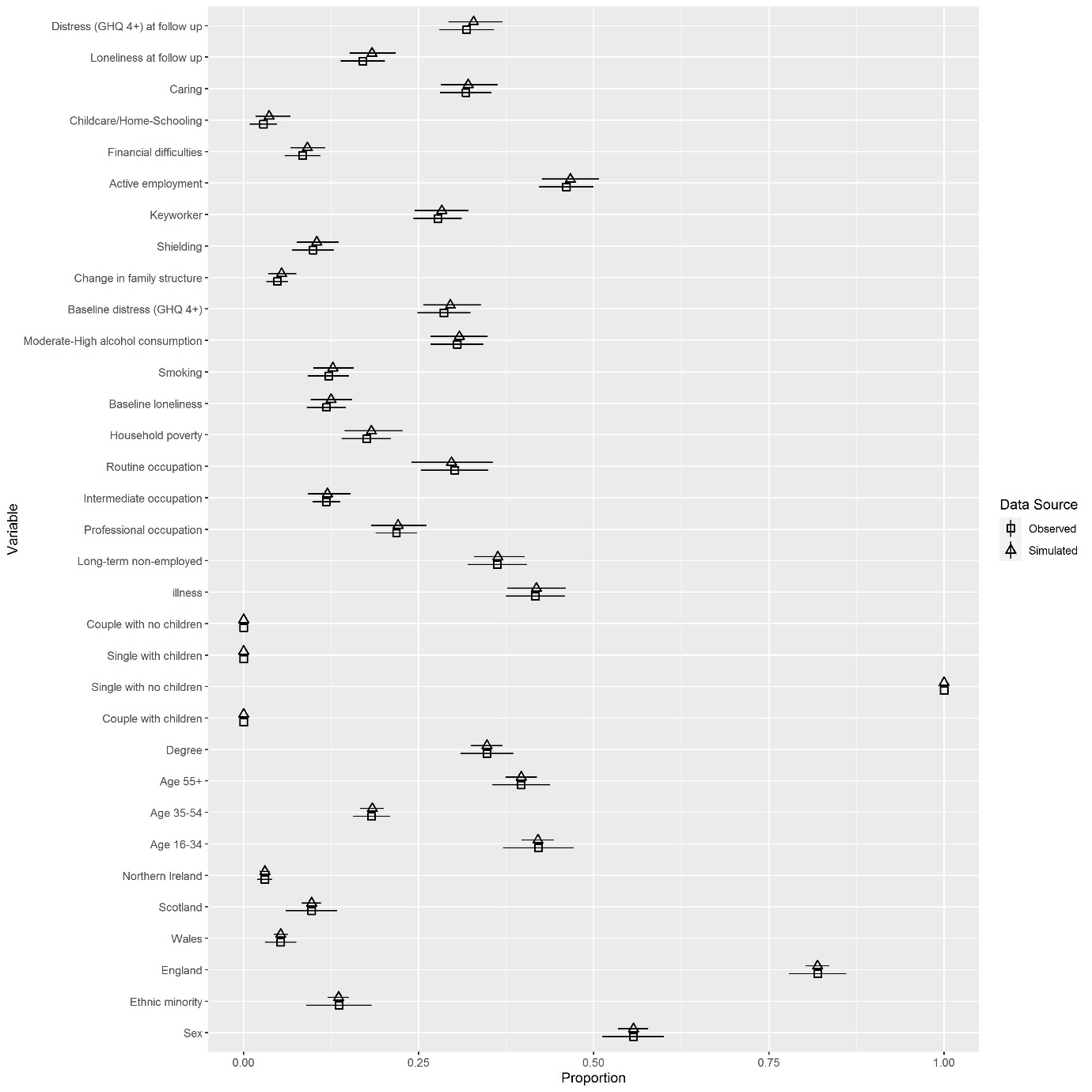

**Figure S13: Validation Comparison of Observed vs Simulated data: January 2021: Single with no children <16 years**

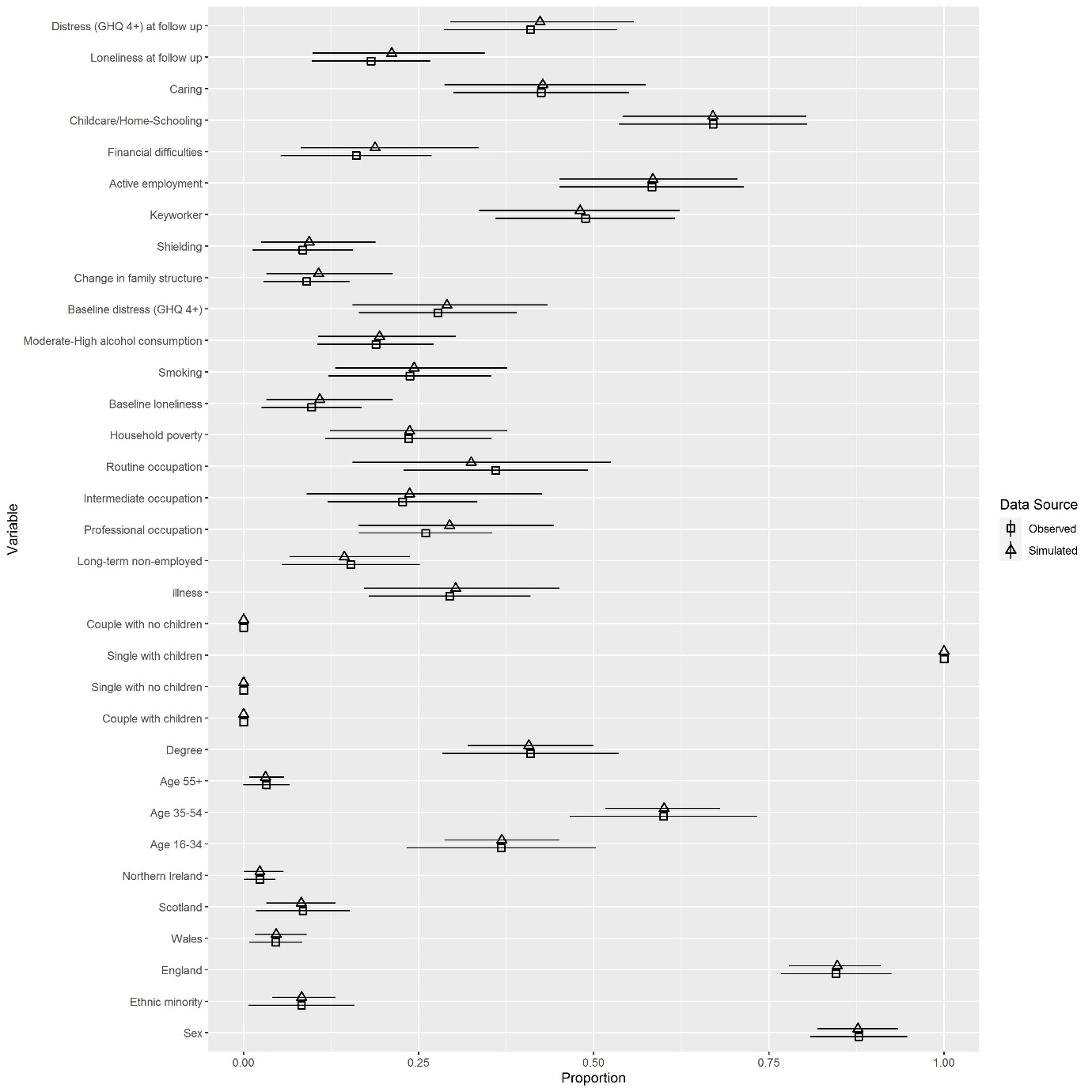

**Figure S14: Validation Comparison of Observed vs Simulated data: January 2021: Single with children <16 years**

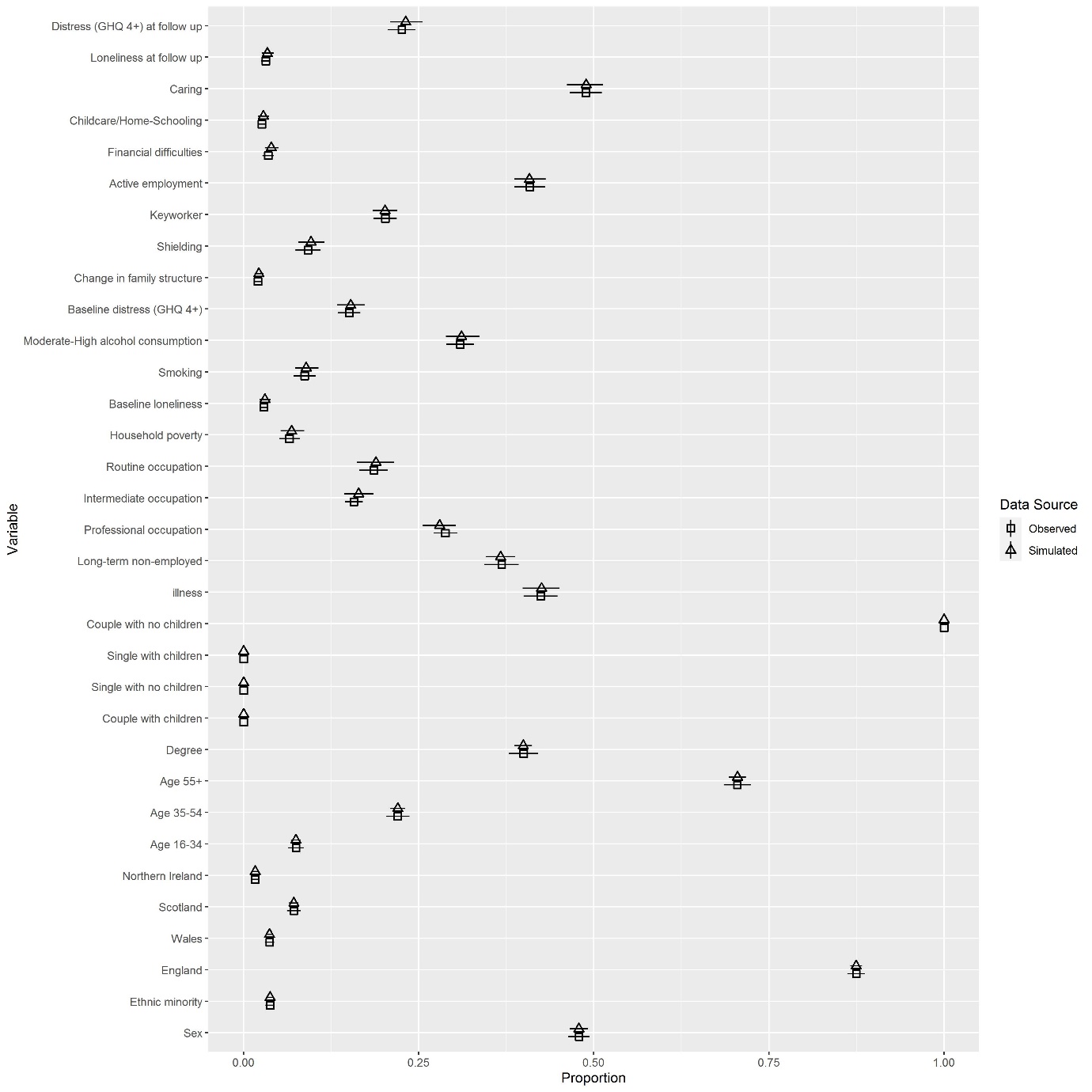

**Figure S15: Validation Comparison of Observed vs Simulated data: January 2021: Couple with no children <16 years**

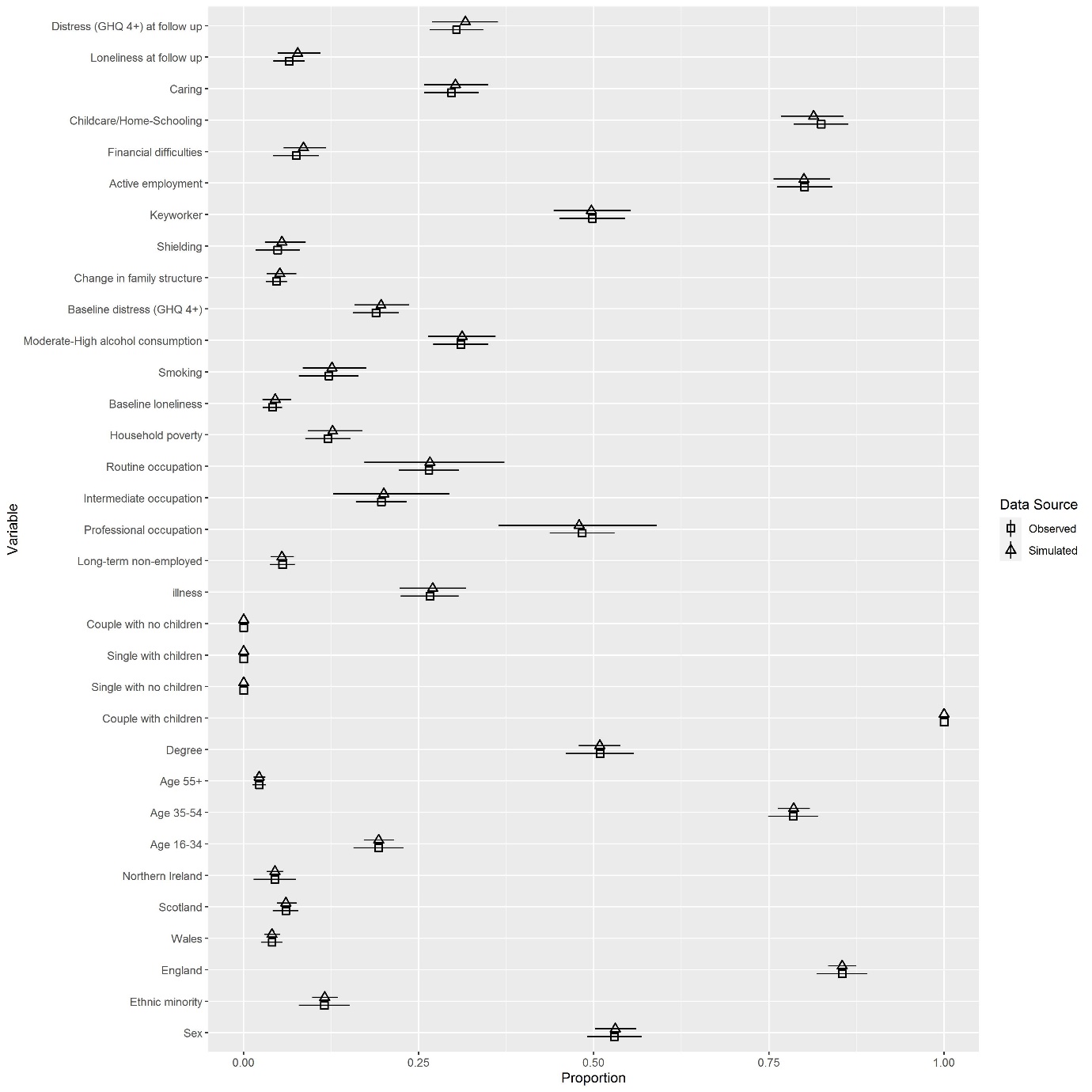

**Figure S16: Validation Comparison of Observed vs Simulated data: January 2021: Couple with children <16 years**

**Table S17: Decomposition details for couples with children <16 years, compared to couples with no children <16 years**

|  | RR | 95% CI  (low) | 95% CI  (high) | Simulated proportion with psychiatric distress for couples with children | Simulated proportion with psychiatric distress for couples with no children |
| --- | --- | --- | --- | --- | --- |
| *April 2020* |  |  |  |  |  |
| *Total Effect* | 1.15 | 0.98 | 1.33 | 0.31 | 0.27 |
| *Active Employment* |  |  |  |  |  |
| Controlled Direct Effect (CDE) | 1.31 | 0.99 | 1.63 |  |  |
| Pure Indirect Effect (PIE) | 1.00 | 0.99 | 1.01 |  |  |
| Reference Interaction (rINT) | 0.85 | 0.65 | 1.06 |  |  |
| Mediated Interaction (mINT) | 0.99 | 0.97 | 1.01 |  |  |
| Portion Eliminated (PE) | 0.84 | 0.62 | 1.06 |  |  |
| *Financial Difficulties* |  |  |  |  |  |
| Controlled Direct Effect (CDE) | 1.12 | 0.95 | 1.29 |  |  |
| Pure Indirect Effect (PIE) | 1.00 | 0.98 | 1.02 |  |  |
| Reference Interaction (rINT) | 1.04 | 1.00 | 1.07 |  |  |
| Mediated Interaction (mINT) | 1.00 | 0.98 | 1.01 |  |  |
| Portion Eliminated (PE) | 1.03 | 0.99 | 1.08 |  |  |
| *Childcare/Home-Schooling* |  |  |  |  |  |
| Controlled Direct Effect (CDE) | 0.94 | 0.67 | 1.22 |  |  |
| Pure Indirect Effect (PIE) | 1.09 | 0.83 | 1.35 |  |  |
| Reference Interaction (rINT) | 1.01 | 0.98 | 1.04 |  |  |
| Mediated Interaction (mINT) | 1.10 | 0.77 | 1.42 |  |  |
| Portion Eliminated (PE) | 1.20 | 0.99 | 1.40 |  |  |
| *Caring* |  |  |  |  |  |
| Controlled Direct Effect (CDE) | 1.05 | 0.84 | 1.25 |  |  |
| Pure Indirect Effect (PIE) | 1.00 | 0.99 | 1.00 |  |  |
| Reference Interaction (rINT) | 1.10 | 0.97 | 1.23 |  |  |
| Mediated Interaction (mINT) | 1.00 | 0.99 | 1.02 |  |  |
| Portion Eliminated (PE) | 1.10 | 0.97 | 1.23 |  |  |
| *Loneliness* |  |  |  |  |  |
| Controlled Direct Effect (CDE) | 1.13 | 0.96 | 1.29 |  |  |
| Pure Indirect Effect (PIE) | 1.01 | 0.99 | 1.04 |  |  |
| Reference Interaction (rINT) | 1.02 | 0.99 | 1.04 |  |  |
| Mediated Interaction (mINT) | 1.00 | 0.99 | 1.01 |  |  |
| Portion Eliminated (PE) | 1.03 | 0.99 | 1.08 |  |  |
| *January 2021* |  |  |  |  |  |
| *Total Effect* | 1.48 | 1.15 | 1.82 | 0.32 | 0.21 |
| *Active Employment* |  |  |  |  |  |
| Controlled Direct Effect (CDE) | 1.45 | 0.95 | 1.94 |  |  |
| Pure Indirect Effect (PIE) | 1.00 | 0.99 | 1.01 |  |  |
| Reference Interaction (rINT) | 1.02 | 0.66 | 1.37 |  |  |
| Mediated Interaction (mINT) | 1.00 | 0.98 | 1.03 |  |  |
| Portion Eliminated (PE) | 1.01 | 0.64 | 1.39 |  |  |
| *Financial Difficulties* |  |  |  |  |  |
| Controlled Direct Effect (CDE) | 1.44 | 1.14 | 1.74 |  |  |
| Pure Indirect Effect (PIE) | 1.01 | 0.99 | 1.02 |  |  |
| Reference Interaction (rINT) | 1.03 | 0.96 | 1.11 |  |  |
| Mediated Interaction (mINT) | 1.01 | 0.98 | 1.04 |  |  |
| Portion Eliminated (PE) | 1.05 | 0.96 | 1.13 |  |  |
| *Childcare/Home-Schooling* |  |  |  |  |  |
| Controlled Direct Effect (CDE) | 1.15 | 0.73 | 1.57 |  |  |
| Pure Indirect Effect (PIE) | 1.31 | 0.95 | 1.67 |  |  |
| Reference Interaction (rINT) | 1.00 | 0.98 | 1.02 |  |  |
| Mediated Interaction (mINT) | 1.00 | 0.54 | 1.47 |  |  |
| Portion Eliminated (PE) | 1.32 | 1.00 | 1.64 |  |  |
| *Caring* |  |  |  |  |  |
| Controlled Direct Effect (CDE) | 1.46 | 1.13 | 1.79 |  |  |
| Pure Indirect Effect (PIE) | 0.99 | 0.98 | 1.01 |  |  |
| Reference Interaction (rINT) | 1.03 | 0.89 | 1.17 |  |  |
| Mediated Interaction (mINT) | 0.99 | 0.96 | 1.02 |  |  |
| Portion Eliminated (PE) | 1.02 | 0.89 | 1.14 |  |  |
| *Loneliness* |  |  |  |  |  |
| Controlled Direct Effect (CDE) | 1.46 | 1.15 | 1.76 |  |  |
| Pure Indirect Effect (PIE) | 1.04 | 0.96 | 1.13 |  |  |
| Reference Interaction (rINT) | 1.03 | 0.97 | 1.08 |  |  |
| Mediated Interaction (mINT) | 1.01 | 0.98 | 1.04 |  |  |
| Portion Eliminated (PE) | 1.08 | 0.97 | 1.20 |  |  |

**Table S18: Decomposition details for couples with children <16 years, compared to couples with no children <16 years: Males**

|  | RR | 95% CI  (low) | 95% CI  (high) | Simulated proportion with psychiatric distress for couples with children | Simulated proportion with psychiatric distress for couples with no children |
| --- | --- | --- | --- | --- | --- |
| *April 2020* |  |  |  |  |  |
| *Total Effect* | 1.36 | 0.95 | 1.77 | 0.26 | 0.19 |
| *Active Employment* |  |  |  |  |  |
| Controlled Direct Effect (CDE) | 1.74 | 0.94 | 2.55 |  |  |
| Pure Indirect Effect (PIE) | 1.00 | 0.98 | 1.02 |  |  |
| Reference Interaction (rINT) | 0.64 | 0.15 | 1.13 |  |  |
| Mediated Interaction (mINT) | 0.97 | 0.90 | 1.03 |  |  |
| Portion Eliminated (PE) | 0.60 | 0.06 | 1.15 |  |  |
| *Financial Difficulties* |  |  |  |  |  |
| Controlled Direct Effect (CDE) | 1.30 | 0.90 | 1.71 |  |  |
| Pure Indirect Effect (PIE) | 0.98 | 0.93 | 1.02 |  |  |
| Reference Interaction (rINT) | 1.12 | 1.00 | 1.23 |  |  |
| Mediated Interaction (mINT) | 0.96 | 0.89 | 1.04 |  |  |
| Portion Eliminated (PE) | 1.06 | 0.95 | 1.17 |  |  |
| *Childcare/Home-Schooling* |  |  |  |  |  |
| Controlled Direct Effect (CDE) | 1.30 | 0.62 | 1.98 |  |  |
| Pure Indirect Effect (PIE) | 1.14 | 0.78 | 1.50 |  |  |
| Reference Interaction (rINT) | 0.99 | 0.93 | 1.06 |  |  |
| Mediated Interaction (mINT) | 0.92 | 0.39 | 1.45 |  |  |
| Portion Eliminated (PE) | 1.05 | 0.62 | 1.49 |  |  |
| *Caring* |  |  |  |  |  |
| Controlled Direct Effect (CDE) | 1.24 | 0.80 | 1.67 |  |  |
| Pure Indirect Effect (PIE) | 1.00 | 0.99 | 1.01 |  |  |
| Reference Interaction (rINT) | 1.12 | 0.86 | 1.38 |  |  |
| Mediated Interaction (mINT) | 1.00 | 0.98 | 1.03 |  |  |
| Portion Eliminated (PE) | 1.12 | 0.86 | 1.39 |  |  |
| *Loneliness* |  |  |  |  |  |
| Controlled Direct Effect (CDE) | 1.33 | 0.93 | 1.73 |  |  |
| Pure Indirect Effect (PIE) | 1.01 | 0.97 | 1.06 |  |  |
| Reference Interaction (rINT) | 1.02 | 0.97 | 1.06 |  |  |
| Mediated Interaction (mINT) | 1.01 | 0.98 | 1.03 |  |  |
| Portion Eliminated (PE) | 1.04 | 0.97 | 1.11 |  |  |
| *January 2021* |  |  |  |  |  |
| *Total Effect* | 1.58 | 0.88 | 2.28 | 0.26 | 0.17 |
| *Active Employment* |  |  |  |  |  |
| Controlled Direct Effect (CDE) | 1.28 | 0.19 | 2.38 |  |  |
| Pure Indirect Effect (PIE) | 1.00 | 0.95 | 1.04 |  |  |
| Reference Interaction (rINT) | 1.25 | 0.43 | 2.06 |  |  |
| Mediated Interaction (mINT) | 1.04 | 0.89 | 1.19 |  |  |
| Portion Eliminated (PE) | 1.28 | 0.33 | 2.22 |  |  |
| *Financial Difficulties* |  |  |  |  |  |
| Controlled Direct Effect (CDE) | 1.56 | 0.94 | 2.19 |  |  |
| Pure Indirect Effect (PIE) | 0.99 | 0.91 | 1.06 |  |  |
| Reference Interaction (rINT) | 1.04 | 0.81 | 1.28 |  |  |
| Mediated Interaction (mINT) | 0.98 | 0.87 | 1.10 |  |  |
| Portion Eliminated (PE) | 1.02 | 0.82 | 1.21 |  |  |
| *Childcare/Home-Schooling* |  |  |  |  |  |
| Controlled Direct Effect (CDE) | 1.33 | 0.55 | 2.10 |  |  |
| Pure Indirect Effect (PIE) | 1.37 | 0.72 | 2.01 |  |  |
| Reference Interaction (rINT) | 1.00 | 0.97 | 1.03 |  |  |
| Mediated Interaction (mINT) | 0.88 | 0.06 | 1.70 |  |  |
| Portion Eliminated (PE) | 1.24 | 0.72 | 1.75 |  |  |
| *Caring* |  |  |  |  |  |
| Controlled Direct Effect (CDE) | 1.66 | 0.95 | 2.37 |  |  |
| Pure Indirect Effect (PIE) | 0.98 | 0.94 | 1.02 |  |  |
| Reference Interaction (rINT) | 0.92 | 0.65 | 1.19 |  |  |
| Mediated Interaction (mINT) | 1.02 | 0.95 | 1.08 |  |  |
| Portion Eliminated (PE) | 0.91 | 0.69 | 1.13 |  |  |
| *Loneliness* |  |  |  |  |  |
| Controlled Direct Effect (CDE) | 1.55 | 0.90 | 2.2 |  |  |
| Pure Indirect Effect (PIE) | 1.00 | 0.85 | 1.15 |  |  |
| Reference Interaction (rINT) | 1.11 | 0.98 | 1.24 |  |  |
| Mediated Interaction (mINT) | 1.00 | 0.87 | 1.12 |  |  |
| Portion Eliminated (PE) | 1.10 | 0.86 | 1.35 |  |  |

**Table S19: Decomposition details for couples with children <16 years, compared to couples with no children <16 years: Females**

|  | RR | 95% CI  (low) | 95% CI  (high) | Simulated proportion with psychiatric distress for couples with children | Simulated proportion with psychiatric distress for couples with no children |
| --- | --- | --- | --- | --- | --- |
| *April 2020* |  |  |  |  |  |
| *Total Effect* | 1.05 | 0.88 | 1.21 | 0.37 | 0.36 |
| *Active Employment* |  |  |  |  |  |
| Controlled Direct Effect (CDE) | 1.08 | 0.84 | 1.32 |  |  |
| Pure Indirect Effect (PIE) | 1.00 | 0.99 | 1.01 |  |  |
| Reference Interaction (rINT) | 0.96 | 0.80 | 1.12 |  |  |
| Mediated Interaction (mINT) | 1.00 | 0.99 | 1.01 |  |  |
| Portion Eliminated (PE) | 0.96 | 0.80 | 1.13 |  |  |
| *Financial Difficulties* |  |  |  |  |  |
| Controlled Direct Effect (CDE) | 1.02 | 0.86 | 1.18 |  |  |
| Pure Indirect Effect (PIE) | 1.01 | 0.99 | 1.04 |  |  |
| Reference Interaction (rINT) | 1.01 | 0.98 | 1.03 |  |  |
| Mediated Interaction (mINT) | 1.00 | 0.99 | 1.02 |  |  |
| Portion Eliminated (PE) | 1.05 | 0.88 | 1.21 |  |  |
| *Childcare/Home-Schooling* |  |  |  |  |  |
| Controlled Direct Effect (CDE) | 0.75 | 0.52 | 0.98 |  |  |
| Pure Indirect Effect (PIE) | 1.06 | 0.73 | 1.39 |  |  |
| Reference Interaction (rINT) | 1.01 | 0.99 | 1.04 |  |  |
| Mediated Interaction (mINT) | 1.22 | 0.83 | 1.61 |  |  |
| Portion Eliminated (PE) | 1.30 | 1.10 | 1.49 |  |  |
| *Caring* |  |  |  |  |  |
| Controlled Direct Effect (CDE) | 0.95 | 0.74 | 1.16 |  |  |
| Pure Indirect Effect (PIE) | 1.00 | 0.99 | 1.01 |  |  |
| Reference Interaction (rINT) | 1.09 | 0.95 | 1.23 |  |  |
| Mediated Interaction (mINT) | 1.01 | 0.99 | 1.02 |  |  |
| Portion Eliminated (PE) | 1.10 | 0.95 | 1.24 |  |  |
| *Loneliness* |  |  |  |  |  |
| Controlled Direct Effect (CDE) | 1.02 | 0.87 | 1.18 |  |  |
| Pure Indirect Effect (PIE) | 1.02 | 0.98 | 1.06 |  |  |
| Reference Interaction (rINT) | 1.01 | 0.99 | 1.04 |  |  |
| Mediated Interaction (mINT) | 1.00 | 0.99 | 1.01 |  |  |
| Portion Eliminated (PE) | 1.03 | 0.98 | 1.08 |  |  |
| *January 2021* |  |  |  |  |  |
| *Total Effect* | 1.45 | 1.08 | 1.83 | 0.37 | 0.26 |
| *Active Employment* |  |  |  |  |  |
| Controlled Direct Effect (CDE) | 1.56 | 1.07 | 2.04 |  |  |
| Pure Indirect Effect (PIE) | 1.00 | 0.99 | 1.01 |  |  |
| Reference Interaction (rINT) | 0.89 | 0.56 | 1.22 |  |  |
| Mediated Interaction (mINT) | 1.00 | 0.98 | 1.03 |  |  |
| Portion Eliminated (PE) | 0.89 | 0.57 | 1.22 |  |  |
| *Financial Difficulties* |  |  |  |  |  |
| Controlled Direct Effect (CDE) | 1.39 | 1.04 | 1.73 |  |  |
| Pure Indirect Effect (PIE) | 1.01 | 0.98 | 1.05 |  |  |
| Reference Interaction (rINT) | 1.02 | 0.98 | 1.07 |  |  |
| Mediated Interaction (mINT) | 1.03 | 0.97 | 1.09 |  |  |
| Portion Eliminated (PE) | 1.07 | 0.98 | 1.16 |  |  |
| *Childcare/Home-Schooling* |  |  |  |  |  |
| Controlled Direct Effect (CDE) | 1.07 | 0.53 | 1.60 |  |  |
| Pure Indirect Effect (PIE) | 1.29 | 0.86 | 1.72 |  |  |
| Reference Interaction (rINT) | 1.00 | 0.97 | 1.04 |  |  |
| Mediated Interaction (mINT) | 1.08 | 0.50 | 1.67 |  |  |
| Portion Eliminated (PE) | 1.38 | 0.95 | 1.81 |  |  |
| *Caring* |  |  |  |  |  |
| Controlled Direct Effect (CDE) | 1.37 | 0.99 | 1.74 |  |  |
| Pure Indirect Effect (PIE) | 1.00 | 0.99 | 1.02 |  |  |
| Reference Interaction (rINT) | 1.10 | 0.94 | 1.26 |  |  |
| Mediated Interaction (mINT) | 0.98 | 0.94 | 1.02 |  |  |
| Portion Eliminated (PE) | 1.09 | 0.95 | 1.23 |  |  |
| *Loneliness* |  |  |  |  |  |
| Controlled Direct Effect (CDE) | 1.42 | 1.07 | 1.77 |  |  |
| Pure Indirect Effect (PIE) | 1.08 | 0.98 | 1.19 |  |  |
| Reference Interaction (rINT) | 0.98 | 0.93 | 1.04 |  |  |
| Mediated Interaction (mINT) | 0.99 | 0.93 | 1.04 |  |  |
| Portion Eliminated (PE) | 1.05 | 0.93 | 1.17 |  |  |

**Table S20: Decomposition details for couples with children <16 years, compared to couples with no children <16 years: Conservative Confounding**

|  | RR | 95% CI  (low) | 95% CI  (high) | Simulated proportion with psychiatric distress for couples with children | Simulated proportion with psychiatric distress for couples with no children |
| --- | --- | --- | --- | --- | --- |
| *April 2020* |  |  |  |  |  |
| *Total Effect* | 1.14 | 0.98 | 1.29 | 0.31 | 0.28 |
| *Active Employment* |  |  |  |  |  |
| Controlled Direct Effect (CDE) | 1.30 | 0.99 | 1.60 |  |  |
| Pure Indirect Effect (PIE) | 1.00 | 0.99 | 1.01 |  |  |
| Reference Interaction (rINT) | 0.85 | 0.64 | 1.06 |  |  |
| Mediated Interaction (mINT) | 0.99 | 0.98 | 1.01 |  |  |
| Portion Eliminated (PE) | 0.84 | 0.62 | 1.06 |  |  |
| *Financial Difficulties* |  |  |  |  |  |
| Controlled Direct Effect (CDE) | 1.11 | 0.96 | 1.26 |  |  |
| Pure Indirect Effect (PIE) | 0.99 | 0.97 | 1.01 |  |  |
| Reference Interaction (rINT) | 1.04 | 1.00 | 1.08 |  |  |
| Mediated Interaction (mINT) | 0.99 | 0.98 | 1.01 |  |  |
| Portion Eliminated (PE) | 1.03 | 0.99 | 1.07 |  |  |
| *Childcare/Home-Schooling* |  |  |  |  |  |
| Controlled Direct Effect (CDE) | 0.93 | 0.67 | 1.18 |  |  |
| Pure Indirect Effect (PIE) | 1.09 | 0.83 | 1.35 |  |  |
| Reference Interaction (rINT) | 1.01 | 0.98 | 1.04 |  |  |
| Mediated Interaction (mINT) | 1.10 | 0.77 | 1.42 |  |  |
| Portion Eliminated (PE) | 1.19 | 0.99 | 1.40 |  |  |
| *Caring* |  |  |  |  |  |
| Controlled Direct Effect (CDE) | 1.03 | 0.84 | 1.23 |  |  |
| Pure Indirect Effect (PIE) | 1.00 | 1.00 | 1.00 |  |  |
| Reference Interaction (rINT) | 1.10 | 0.97 | 1.23 |  |  |
| Mediated Interaction (mINT) | 1.00 | 0.99 | 1.01 |  |  |
| Portion Eliminated (PE) | 1.10 | 0.97 | 1.23 |  |  |
| *Loneliness* |  |  |  |  |  |
| Controlled Direct Effect (CDE) | 1.11 | 0.96 | 1.26 |  |  |
| Pure Indirect Effect (PIE) | 1.01 | 0.99 | 1.04 |  |  |
| Reference Interaction (rINT) | 1.02 | 0.99 | 1.04 |  |  |
| Mediated Interaction (mINT) | 1.00 | 0.99 | 1.01 |  |  |
| Portion Eliminated (PE) | 1.03 | 0.99 | 1.07 |  |  |
| *January 2021* |  |  |  |  |  |
| *Total Effect* | 1.49 | 1.20 | 1.78 | 0.31 | 0.21 |
| *Active Employment* |  |  |  |  |  |
| Controlled Direct Effect (CDE) | 1.45 | 0.95 | 1.94 |  |  |
| Pure Indirect Effect (PIE) | 1.00 | 0.99 | 1.01 |  |  |
| Reference Interaction (rINT) | 1.02 | 0.64 | 1.39 |  |  |
| Mediated Interaction (mINT) | 1.00 | 0.98 | 1.02 |  |  |
| Portion Eliminated (PE) | 1.02 | 0.63 | 1.40 |  |  |
| *Financial Difficulties* |  |  |  |  |  |
| Controlled Direct Effect (CDE) | 1.45 | 1.17 | 1.72 |  |  |
| Pure Indirect Effect (PIE) | 1.00 | 0.99 | 1.02 |  |  |
| Reference Interaction (rINT) | 1.04 | 0.96 | 1.11 |  |  |
| Mediated Interaction (mINT) | 1.00 | 0.98 | 1.03 |  |  |
| Portion Eliminated (PE) | 1.04 | 0.96 | 1.13 |  |  |
| *Childcare/Home-Schooling* |  |  |  |  |  |
| Controlled Direct Effect (CDE) | 1.16 | 0.75 | 1.56 |  |  |
| Pure Indirect Effect (PIE) | 1.32 | 0.95 | 1.68 |  |  |
| Reference Interaction (rINT) | 1.00 | 0.98 | 1.02 |  |  |
| Mediated Interaction (mINT) | 1.00 | 0.53 | 1.47 |  |  |
| Portion Eliminated (PE) | 1.32 | 0.99 | 1.65 |  |  |
| *Caring* |  |  |  |  |  |
| Controlled Direct Effect (CDE) | 1.46 | 1.16 | 1.77 |  |  |
| Pure Indirect Effect (PIE) | 0.99 | 0.98 | 1.01 |  |  |
| Reference Interaction (rINT) | 1.03 | 0.89 | 1.18 |  |  |
| Mediated Interaction (mINT) | 0.99 | 0.96 | 1.02 |  |  |
| Portion Eliminated (PE) | 1.02 | 0.90 | 1.14 |  |  |
| *Loneliness* |  |  |  |  |  |
| Controlled Direct Effect (CDE) | 1.46 | 1.17 | 1.74 |  |  |
| Pure Indirect Effect (PIE) | 1.04 | 0.97 | 1.11 |  |  |
| Reference Interaction (rINT) | 1.03 | 0.97 | 1.08 |  |  |
| Mediated Interaction (mINT) | 1.01 | 0.98 | 1.03 |  |  |
| Portion Eliminated (PE) | 1.07 | 0.97 | 1.17 |  |  |

**Table S21: Decomposition details for singles with no children <16 years, compared to couples with no children <16 years**

|  | RR | 95% CI  (low) | 95% CI  (high) | Simulated proportion with psychiatric distress for singles with no children | Simulated proportion with psychiatric distress for couples with no children |
| --- | --- | --- | --- | --- | --- |
| *April 2020* |  |  |  |  |  |
| *Total Effect* | 1.28 | 1.11 | 1.44 | 0.37 | 0.29 |
| *Active Employment* |  |  |  |  |  |
| Controlled Direct Effect (CDE) | 1.26 | 1.05 | 1.46 |  |  |
| Pure Indirect Effect (PIE) | 1.00 | 0.98 | 1.02 |  |  |
| Reference Interaction (rINT) | 1.02 | 0.91 | 1.12 |  |  |
| Mediated Interaction (mINT) | 1.00 | 0.98 | 1.02 |  |  |
| Portion Eliminated (PE) | 1.01 | 0.92 | 1.11 |  |  |
| *Financial Difficulties* |  |  |  |  |  |
| Controlled Direct Effect (CDE) | 1.21 | 1.05 | 1.37 |  |  |
| Pure Indirect Effect (PIE) | 1.05 | 1.01 | 1.08 |  |  |
| Reference Interaction (rINT) | 1.01 | 0.98 | 1.03 |  |  |
| Mediated Interaction (mINT) | 1.01 | 0.98 | 1.04 |  |  |
| Portion Eliminated (PE) | 1.07 | 1.02 | 1.12 |  |  |
| *Childcare/Home-Schooling* |  |  |  |  |  |
| Controlled Direct Effect (CDE) | 1.28 | 1.12 | 1.44 |  |  |
| Pure Indirect Effect (PIE) | 1.00 | 0.99 | 1.01 |  |  |
| Reference Interaction (rINT) | 1.01 | 0.98 | 1.04 |  |  |
| Mediated Interaction (mINT) | 1.00 | 0.99 | 1.01 |  |  |
| Portion Eliminated (PE) | 1.00 | 0.98 | 1.03 |  |  |
| *Caring* |  |  |  |  |  |
| Controlled Direct Effect (CDE) | 1.26 | 1.06 | 1.46 |  |  |
| Pure Indirect Effect (PIE) | 1.00 | 0.99 | 1.01 |  |  |
| Reference Interaction (rINT) | 1.02 | 0.92 | 1.12 |  |  |
| Mediated Interaction (mINT) | 1.00 | 0.98 | 1.01 |  |  |
| Portion Eliminated (PE) | 1.02 | 0.92 | 1.11 |  |  |
| *Loneliness* |  |  |  |  |  |
| Controlled Direct Effect (CDE) | 1.11 | 0.97 | 1.26 |  |  |
| Pure Indirect Effect (PIE) | 1.17 | 1.11 | 1.24 |  |  |
| Reference Interaction (rINT) | 1.00 | 0.98 | 1.02 |  |  |
| Mediated Interaction (mINT) | 1.00 | 0.95 | 1.06 |  |  |
| Portion Eliminated (PE) | 1.18 | 1.12 | 1.24 |  |  |
| *January 2021* |  |  |  |  |  |
| *Total Effect* | 1.55 | 1.27 | 1.83 | 0.33 | 0.21 |
| *Active Employment* |  |  |  |  |  |
| Controlled Direct Effect (CDE) | 1.46 | 1.16 | 1.76 |  |  |
| Pure Indirect Effect (PIE) | 1.00 | 0.98 | 1.02 |  |  |
| Reference Interaction (rINT) | 1.10 | 0.93 | 1.28 |  |  |
| Mediated Interaction (mINT) | 0.98 | 0.95 | 1.01 |  |  |
| Portion Eliminated (PE) | 1.09 | 0.93 | 1.24 |  |  |
| *Financial Difficulties* |  |  |  |  |  |
| Controlled Direct Effect (CDE) | 1.50 | 1.25 | 1.75 |  |  |
| Pure Indirect Effect (PIE) | 1.01 | 0.98 | 1.04 |  |  |
| Reference Interaction (rINT) | 1.02 | 0.98 | 1.07 |  |  |
| Mediated Interaction (mINT) | 1.02 | 0.98 | 1.06 |  |  |
| Portion Eliminated (PE) | 1.05 | 0.99 | 1.12 |  |  |
| *Childcare/Home-Schooling* |  |  |  |  |  |
| Controlled Direct Effect (CDE) | 1.57 | 1.30 | 1.83 |  |  |
| Pure Indirect Effect (PIE) | 1.00 | 0.99 | 1.01 |  |  |
| Reference Interaction (rINT) | 0.98 | 0.96 | 1.01 |  |  |
| Mediated Interaction (mINT) | 1.00 | 0.98 | 1.01 |  |  |
| Portion Eliminated (PE) | 0.98 | 0.95 | 1.02 |  |  |
| *Caring* |  |  |  |  |  |
| Controlled Direct Effect (CDE) | 1.55 | 1.26 | 1.84 |  |  |
| Pure Indirect Effect (PIE) | 1.00 | 0.99 | 1.01 |  |  |
| Reference Interaction (rINT) | 1.01 | 0.91 | 1.12 |  |  |
| Mediated Interaction (mINT) | 1.00 | 0.99 | 1.01 |  |  |
| Portion Eliminated (PE) | 1.01 | 0.91 | 1.12 |  |  |
| *Loneliness* |  |  |  |  |  |
| Controlled Direct Effect (CDE) | 1.41 | 1.16 | 1.66 |  |  |
| Pure Indirect Effect (PIE) | 1.28 | 1.16 | 1.40 |  |  |
| Reference Interaction (rINT) | 0.97 | 0.92 | 1.01 |  |  |
| Mediated Interaction (mINT) | 0.91 | 0.81 | 1.02 |  |  |
| Portion Eliminated (PE) | 1.16 | 1.05 | 1.27 |  |  |

**Table S22: Decomposition details for singles with no children <16 years, compared to couples with no children <16 years: Males**

|  | RR | 95% CI  (low) | 95% CI  (high) | Simulated proportion with psychiatric distress for singles with no children | Simulated proportion with psychiatric distress for couples with no children |
| --- | --- | --- | --- | --- | --- |
| *April 2020* |  |  |  |  |  |
| *Total Effect* | 1.45 | 1.09 | 1.81 | 0.31 | 0.22 |
| *Active Employment* |  |  |  |  |  |
| Controlled Direct Effect (CDE) | 1.33 | 0.93 | 1.72 |  |  |
| Pure Indirect Effect (PIE) | 1.00 | 0.97 | 1.02 |  |  |
| Reference Interaction (rINT) | 1.15 | 0.92 | 1.37 |  |  |
| Mediated Interaction (mINT) | 0.98 | 0.94 | 1.02 |  |  |
| Portion Eliminated (PE) | 1.12 | 0.92 | 1.32 |  |  |
| *Financial Difficulties* |  |  |  |  |  |
| Controlled Direct Effect (CDE) | 1.33 | 1.01 | 1.65 |  |  |
| Pure Indirect Effect (PIE) | 1.05 | 0.98 | 1.12 |  |  |
| Reference Interaction (rINT) | 1.04 | 0.97 | 1.11 |  |  |
| Mediated Interaction (mINT) | 1.04 | 0.96 | 1.12 |  |  |
| Portion Eliminated (PE) | 1.13 | 1.00 | 1.25 |  |  |
| *Childcare/Home-Schooling* |  |  |  |  |  |
| Controlled Direct Effect (CDE) | 1.45 | 1.11 | 1.80 |  |  |
| Pure Indirect Effect (PIE) | 1.00 | 0.98 | 1.01 |  |  |
| Reference Interaction (rINT) | 1.00 | 0.95 | 1.05 |  |  |
| Mediated Interaction (mINT) | 1.00 | 0.98 | 1.02 |  |  |
| Portion Eliminated (PE) | 1.00 | 0.95 | 1.05 |  |  |
| *Caring* |  |  |  |  |  |
| Controlled Direct Effect (CDE) | 1.44 | 1.03 | 1.84 |  |  |
| Pure Indirect Effect (PIE) | 1.00 | 0.98 | 1.02 |  |  |
| Reference Interaction (rINT) | 1.02 | 0.83 | 1.20 |  |  |
| Mediated Interaction (mINT) | 1.00 | 0.96 | 1.03 |  |  |
| Portion Eliminated (PE) | 1.01 | 0.85 | 1.18 |  |  |
| *Loneliness* |  |  |  |  |  |
| Controlled Direct Effect (CDE) | 1.18 | 0.88 | 1.48 |  |  |
| Pure Indirect Effect (PIE) | 1.19 | 1.04 | 1.34 |  |  |
| Reference Interaction (rINT) | 1.02 | 0.98 | 1.05 |  |  |
| Mediated Interaction (mINT) | 1.09 | 0.93 | 1.24 |  |  |
| Portion Eliminated (PE) | 1.29 | 1.15 | 1.43 |  |  |
| *January 2021* |  |  |  |  |  |
| *Total Effect* | 1.58 | 1.02 | 2.15 | 0.26 | 0.17 |
| *Active Employment* |  |  |  |  |  |
| Controlled Direct Effect (CDE) | 1.30 | 0.76 | 1.85 |  |  |
| Pure Indirect Effect (PIE) | 1.00 | 0.96 | 1.04 |  |  |
| Reference Interaction (rINT) | 1.34 | 1.00 | 1.67 |  |  |
| Mediated Interaction (mINT) | 0.94 | 0.86 | 1.02 |  |  |
| Portion Eliminated (PE) | 1.28 | 0.99 | 1.57 |  |  |
| *Financial Difficulties* |  |  |  |  |  |
| Controlled Direct Effect (CDE) | 1.51 | 1.02 | 2.00 |  |  |
| Pure Indirect Effect (PIE) | 1.01 | 0.96 | 1.06 |  |  |
| Reference Interaction (rINT) | 1.05 | 0.90 | 1.20 |  |  |
| Mediated Interaction (mINT) | 1.01 | 0.94 | 1.09 |  |  |
| Portion Eliminated (PE) | 1.07 | 0.91 | 1.23 |  |  |
| *Childcare/Home-Schooling* |  |  |  |  |  |
| Controlled Direct Effect (CDE) | 1.61 | 1.09 | 2.14 |  |  |
| Pure Indirect Effect (PIE) | 1.01 | 0.98 | 1.04 |  |  |
| Reference Interaction (rINT) | 0.97 | 0.91 | 1.04 |  |  |
| Mediated Interaction (mINT) | 0.99 | 0.93 | 1.05 |  |  |
| Portion Eliminated (PE) | 0.97 | 0.88 | 1.07 |  |  |
| *Caring* |  |  |  |  |  |
| Controlled Direct Effect (CDE) | 1.49 | 0.97 | 2.02 |  |  |
| Pure Indirect Effect (PIE) | 1.00 | 0.97 | 1.02 |  |  |
| Reference Interaction (rINT) | 1.09 | 0.92 | 1.26 |  |  |
| Mediated Interaction (mINT) | 1.00 | 0.97 | 1.03 |  |  |
| Portion Eliminated (PE) | 1.09 | 0.91 | 1.26 |  |  |
| *Loneliness* |  |  |  |  |  |
| Controlled Direct Effect (CDE) | 1.45 | 0.94 | 1.96 |  |  |
| Pure Indirect Effect (PIE) | 1.35 | 1.07 | 1.63 |  |  |
| Reference Interaction (rINT) | 0.93 | 0.82 | 1.03 |  |  |
| Mediated Interaction (mINT) | 0.83 | 0.56 | 1.09 |  |  |
| Portion Eliminated (PE) | 1.10 | 0.87 | 1.34 |  |  |

**Table S23: Decomposition details for singles with no children <16 years, compared to couples with no children <16 years: Females**

|  | RR | 95% CI  (low) | 95% CI  (high) | Simulated proportion with psychiatric distress for singles with no children | Simulated proportion with psychiatric distress for couples with no children |
| --- | --- | --- | --- | --- | --- |
| *April 2020* |  |  |  |  |  |
| *Total Effect* | 1.19 | 1.02 | 1.37 | 0.41 | 0.34 |
| *Active Employment* |  |  |  |  |  |
| Controlled Direct Effect (CDE) | 1.23 | 1.01 | 1.46 |  |  |
| Pure Indirect Effect (PIE) | 1.00 | 0.98 | 1.02 |  |  |
| Reference Interaction (rINT) | 0.96 | 0.84 | 1.07 |  |  |
| Mediated Interaction (mINT) | 1.01 | 0.99 | 1.03 |  |  |
| Portion Eliminated (PE) | 0.97 | 0.86 | 1.07 |  |  |
| *Financial Difficulties* |  |  |  |  |  |
| Controlled Direct Effect (CDE) | 1.15 | 0.98 | 1.33 |  |  |
| Pure Indirect Effect (PIE) | 1.05 | 1.01 | 1.08 |  |  |
| Reference Interaction (rINT) | 1.00 | 0.98 | 1.02 |  |  |
| Mediated Interaction (mINT) | 1.00 | 0.96 | 1.03 |  |  |
| Portion Eliminated (PE) | 1.04 | 1.00 | 1.09 |  |  |
| *Childcare/Home-Schooling* |  |  |  |  |  |
| Controlled Direct Effect (CDE) | 1.19 | 1.02 | 1.36 |  |  |
| Pure Indirect Effect (PIE) | 1.00 | 0.99 | 1.01 |  |  |
| Reference Interaction (rINT) | 1.01 | 0.97 | 1.04 |  |  |
| Mediated Interaction (mINT) | 1.00 | 0.98 | 1.01 |  |  |
| Portion Eliminated (PE) | 1.00 | 0.98 | 1.03 |  |  |
| *Caring* |  |  |  |  |  |
| Controlled Direct Effect (CDE) | 1.17 | 0.96 | 1.38 |  |  |
| Pure Indirect Effect (PIE) | 1.00 | 0.99 | 1.01 |  |  |
| Reference Interaction (rINT) | 1.02 | 0.90 | 1.14 |  |  |
| Mediated Interaction (mINT) | 1.00 | 0.98 | 1.01 |  |  |
| Portion Eliminated (PE) | 1.02 | 0.91 | 1.13 |  |  |
| *Loneliness* |  |  |  |  |  |
| Controlled Direct Effect (CDE) | 1.09 | 0.92 | 1.25 |  |  |
| Pure Indirect Effect (PIE) | 1.17 | 1.10 | 1.23 |  |  |
| Reference Interaction (rINT) | 0.98 | 0.96 | 1.01 |  |  |
| Mediated Interaction (mINT) | 0.97 | 0.92 | 1.01 |  |  |
| Portion Eliminated (PE) | 1.12 | 1.05 | 1.19 |  |  |
| *January 2021* |  |  |  |  |  |
| *Total Effect* | 1.55 | 1.23 | 1.87 | 0.38 | 0.25 |
| *Active Employment* |  |  |  |  |  |
| Controlled Direct Effect (CDE) | 1.55 | 1.20 | 1.91 |  |  |
| Pure Indirect Effect (PIE) | 1.00 | 0.98 | 1.02 |  |  |
| Reference Interaction (rINT) | 0.99 | 0.80 | 1.18 |  |  |
| Mediated Interaction (mINT) | 1.00 | 0.97 | 1.03 |  |  |
| Portion Eliminated (PE) | 0.99 | 0.82 | 1.17 |  |  |
| *Financial Difficulties* |  |  |  |  |  |
| Controlled Direct Effect (CDE) | 1.50 | 1.20 | 1.80 |  |  |
| Pure Indirect Effect (PIE) | 1.01 | 0.98 | 1.05 |  |  |
| Reference Interaction (rINT) | 1.01 | 0.99 | 1.03 |  |  |
| Mediated Interaction (mINT) | 1.02 | 0.97 | 1.07 |  |  |
| Portion Eliminated (PE) | 1.05 | 0.99 | 1.10 |  |  |
| *Childcare/Home-Schooling* |  |  |  |  |  |
| Controlled Direct Effect (CDE) | 1.56 | 1.25 | 1.86 |  |  |
| Pure Indirect Effect (PIE) | 1.00 | 0.99 | 1.01 |  |  |
| Reference Interaction (rINT) | 0.99 | 0.96 | 1.02 |  |  |
| Mediated Interaction (mINT) | 1.00 | 0.99 | 1.02 |  |  |
| Portion Eliminated (PE) | 0.99 | 0.96 | 1.02 |  |  |
| *Caring* |  |  |  |  |  |
| Controlled Direct Effect (CDE) | 1.59 | 1.23 | 1.95 |  |  |
| Pure Indirect Effect (PIE) | 1.00 | 0.99 | 1.01 |  |  |
| Reference Interaction (rINT) | 0.96 | 0.83 | 1.09 |  |  |
| Mediated Interaction (mINT) | 1.00 | 0.99 | 1.01 |  |  |
| Portion Eliminated (PE) | 0.96 | 0.82 | 1.09 |  |  |
| *Loneliness* |  |  |  |  |  |
| Controlled Direct Effect (CDE) | 1.40 | 1.11 | 1.70 |  |  |
| Pure Indirect Effect (PIE) | 1.24 | 1.12 | 1.36 |  |  |
| Reference Interaction (rINT) | 0.98 | 0.95 | 1.02 |  |  |
| Mediated Interaction (mINT) | 0.95 | 0.86 | 1.04 |  |  |
| Portion Eliminated (PE) | 1.18 | 1.07 | 1.29 |  |  |

**Table S24: Decomposition details for singles with no children <16 years, compared to couples with no children <16 years: Conservative Confounding**

|  | RR | 95% CI  (low) | 95% CI  (high) | Simulated proportion with psychiatric distress for singles with no children | Simulated proportion with psychiatric distress for couples with no children |
| --- | --- | --- | --- | --- | --- |
| *April 2020* |  |  |  |  |  |
| *Total Effect* | 1.13 | 1.00 | 1.26 | 0.36 | 0.32 |
| *Active Employment* |  |  |  |  |  |
| Controlled Direct Effect (CDE) | 1.12 | 0.95 | 1.28 |  |  |
| Pure Indirect Effect (PIE) | 1.00 | 1.00 | 1.00 |  |  |
| Reference Interaction (rINT) | 1.01 | 0.93 | 1.10 |  |  |
| Mediated Interaction (mINT) | 1.00 | 0.99 | 1.00 |  |  |
| Portion Eliminated (PE) | 1.01 | 0.93 | 1.10 |  |  |
| *Financial Difficulties* |  |  |  |  |  |
| Controlled Direct Effect (CDE) | 1.09 | 0.97 | 1.21 |  |  |
| Pure Indirect Effect (PIE) | 1.03 | 1.00 | 1.05 |  |  |
| Reference Interaction (rINT) | 1.01 | 0.98 | 1.04 |  |  |
| Mediated Interaction (mINT) | 1.01 | 0.99 | 1.03 |  |  |
| Portion Eliminated (PE) | 1.05 | 1.00 | 1.09 |  |  |
| *Childcare/Home-Schooling* |  |  |  |  |  |
| Controlled Direct Effect (CDE) | 1.13 | 1.01 | 1.26 |  |  |
| Pure Indirect Effect (PIE) | 1.00 | 0.99 | 1.00 |  |  |
| Reference Interaction (rINT) | 1.00 | 0.98 | 1.03 |  |  |
| Mediated Interaction (mINT) | 1.00 | 0.99 | 1.01 |  |  |
| Portion Eliminated (PE) | 1.00 | 0.98 | 1.02 |  |  |
| *Caring* |  |  |  |  |  |
| Controlled Direct Effect (CDE) | 1.12 | 0.95 | 1.28 |  |  |
| Pure Indirect Effect (PIE) | 1.00 | 0.99 | 1.01 |  |  |
| Reference Interaction (rINT) | 1.02 | 0.93 | 1.11 |  |  |
| Mediated Interaction (mINT) | 1.00 | 0.99 | 1.01 |  |  |
| Portion Eliminated (PE) | 1.02 | 0.93 | 1.10 |  |  |
| *Loneliness* |  |  |  |  |  |
| Controlled Direct Effect (CDE) | 1.01 | 0.89 | 1.13 |  |  |
| Pure Indirect Effect (PIE) | 1.12 | 1.07 | 1.17 |  |  |
| Reference Interaction (rINT) | 1.00 | 0.98 | 1.03 |  |  |
| Mediated Interaction (mINT) | 1.01 | 0.97 | 1.05 |  |  |
| Portion Eliminated (PE) | 1.13 | 1.08 | 1.19 |  |  |
| *January 2021* |  |  |  |  |  |
| *Total Effect* | 1.16 | 1.00 | 1.32 | 0.32 | 0.28 |
| *Active Employment* |  |  |  |  |  |
| Controlled Direct Effect (CDE) | 1.09 | 0.89 | 1.29 |  |  |
| Pure Indirect Effect (PIE) | 1.00 | 1.00 | 1.00 |  |  |
| Reference Interaction (rINT) | 1.07 | 0.95 | 1.18 |  |  |
| Mediated Interaction (mINT) | 1.00 | 0.99 | 1.01 |  |  |
| Portion Eliminated (PE) | 1.06 | 0.95 | 1.18 |  |  |
| *Financial Difficulties* |  |  |  |  |  |
| Controlled Direct Effect (CDE) | 1.12 | 0.98 | 1.27 |  |  |
| Pure Indirect Effect (PIE) | 1.00 | 0.99 | 1.02 |  |  |
| Reference Interaction (rINT) | 1.03 | 0.97 | 1.08 |  |  |
| Mediated Interaction (mINT) | 1.00 | 0.99 | 1.02 |  |  |
| Portion Eliminated (PE) | 1.03 | 0.97 | 1.09 |  |  |
| *Childcare/Home-Schooling* |  |  |  |  |  |
| Controlled Direct Effect (CDE) | 1.17 | 1.01 | 1.33 |  |  |
| Pure Indirect Effect (PIE) | 1.00 | 0.99 | 1.01 |  |  |
| Reference Interaction (rINT) | 0.99 | 0.96 | 1.01 |  |  |
| Mediated Interaction (mINT) | 1.00 | 0.99 | 1.01 |  |  |
| Portion Eliminated (PE) | 0.99 | 0.96 | 1.01 |  |  |
| *Caring* |  |  |  |  |  |
| Controlled Direct Effect (CDE) | 1.16 | 0.97 | 1.34 |  |  |
| Pure Indirect Effect (PIE) | 1.00 | 1.00 | 1.00 |  |  |
| Reference Interaction (rINT) | 1.01 | 0.92 | 1.09 |  |  |
| Mediated Interaction (mINT) | 1.00 | 0.99 | 1.01 |  |  |
| Portion Eliminated (PE) | 1.00 | 0.92 | 1.09 |  |  |
| *Loneliness* |  |  |  |  |  |
| Controlled Direct Effect (CDE) | 1.11 | 0.94 | 1.28 |  |  |
| Pure Indirect Effect (PIE) | 1.12 | 1.06 | 1.19 |  |  |
| Reference Interaction (rINT) | 0.95 | 0.90 | 1.01 |  |  |
| Mediated Interaction (mINT) | 0.96 | 0.92 | 1.01 |  |  |
| Portion Eliminated (PE) | 1.04 | 0.96 | 1.13 |  |  |

**Table S25: Decomposition details for singles with children <16 years, compared to couples with children <16 years**

|  | RR | 95% CI  (low) | 95% CI  (high) | Simulated proportion with psychiatric distress for singles with children | Simulated proportion with psychiatric distress for couples with children |
| --- | --- | --- | --- | --- | --- |
| *April 2020* |  |  |  |  |  |
| *Total Effect* | 1.41 | 1.03 | 1.79 | 0.54 | 0.39 |
| *Active Employment* |  |  |  |  |  |
| Controlled Direct Effect (CDE) | 1.39 | 0.93 | 1.85 |  |  |
| Pure Indirect Effect (PIE) | 1.01 | 0.98 | 1.04 |  |  |
| Reference Interaction (rINT) | 1.00 | 0.66 | 1.35 |  |  |
| Mediated Interaction (mINT) | 1.00 | 0.93 | 1.07 |  |  |
| Portion Eliminated (PE) | 1.01 | 0.72 | 1.31 |  |  |
| *Financial Difficulties* |  |  |  |  |  |
| Controlled Direct Effect (CDE) | 1.41 | 1.05 | 1.77 |  |  |
| Pure Indirect Effect (PIE) | 1.03 | 0.97 | 1.08 |  |  |
| Reference Interaction (rINT) | 0.98 | 0.93 | 1.03 |  |  |
| Mediated Interaction (mINT) | 0.99 | 0.95 | 1.03 |  |  |
| Portion Eliminated (PE) | 0.99 | 0.92 | 1.07 |  |  |
| *Childcare/Home-Schooling* |  |  |  |  |  |
| Controlled Direct Effect (CDE) | 1.69 | 1.09 | 2.29 |  |  |
| Pure Indirect Effect (PIE) | 0.99 | 0.96 | 1.02 |  |  |
| Reference Interaction (rINT) | 0.71 | 0.19 | 1.23 |  |  |
| Mediated Interaction (mINT) | 1.01 | 0.96 | 1.05 |  |  |
| Portion Eliminated (PE) | 0.71 | 0.21 | 1.22 |  |  |
| *Caring* |  |  |  |  |  |
| Controlled Direct Effect (CDE) | 1.21 | 0.80 | 1.63 |  |  |
| Pure Indirect Effect (PIE) | 1.01 | 0.99 | 1.03 |  |  |
| Reference Interaction (rINT) | 1.16 | 0.87 | 1.46 |  |  |
| Mediated Interaction (mINT) | 1.02 | 0.96 | 1.07 |  |  |
| Portion Eliminated (PE) | 1.19 | 0.86 | 1.52 |  |  |
| *Loneliness* |  |  |  |  |  |
| Controlled Direct Effect (CDE) | 1.33 | 0.96 | 1.70 |  |  |
| Pure Indirect Effect (PIE) | 1.15 | 1.01 | 1.28 |  |  |
| Reference Interaction (rINT) | 0.98 | 0.93 | 1.02 |  |  |
| Mediated Interaction (mINT) | 0.97 | 0.90 | 1.04 |  |  |
| Portion Eliminated (PE) | 1.09 | 0.96 | 1.23 |  |  |
| *January 2021* |  |  |  |  |  |
| *Total Effect* | 1.22 | 0.76 | 1.69 | 0.43 | 0.35 |
| *Active Employment* |  |  |  |  |  |
| Controlled Direct Effect (CDE) | 1.09 | 0.54 | 1.64 |  |  |
| Pure Indirect Effect (PIE) | 1.01 | 0.96 | 1.06 |  |  |
| Reference Interaction (rINT) | 1.18 | 0.72 | 1.64 |  |  |
| Mediated Interaction (mINT) | 0.97 | 0.86 | 1.07 |  |  |
| Portion Eliminated (PE) | 1.16 | 0.76 | 1.55 |  |  |
| *Financial Difficulties* |  |  |  |  |  |
| Controlled Direct Effect (CDE) | 1.29 | 0.81 | 1.77 |  |  |
| Pure Indirect Effect (PIE) | 1.06 | 0.95 | 1.17 |  |  |
| Reference Interaction (rINT) | 0.94 | 0.86 | 1.02 |  |  |
| Mediated Interaction (mINT) | 0.92 | 0.77 | 1.07 |  |  |
| Portion Eliminated (PE) | 0.92 | 0.79 | 1.06 |  |  |
| *Childcare/Home-Schooling* |  |  |  |  |  |
| Controlled Direct Effect (CDE) | 1.32 | 0.70 | 1.94 |  |  |
| Pure Indirect Effect (PIE) | 0.94 | 0.85 | 1.03 |  |  |
| Reference Interaction (rINT) | 0.97 | 0.40 | 1.54 |  |  |
| Mediated Interaction (mINT) | 1.01 | 0.88 | 1.14 |  |  |
| Portion Eliminated (PE) | 0.92 | 0.43 | 1.42 |  |  |
| *Caring* |  |  |  |  |  |
| Controlled Direct Effect (CDE) | 1.50 | 0.95 | 2.04 |  |  |
| Pure Indirect Effect (PIE) | 1.02 | 0.97 | 1.07 |  |  |
| Reference Interaction (rINT) | 0.79 | 0.61 | 0.98 |  |  |
| Mediated Interaction (mINT) | 0.90 | 0.73 | 1.06 |  |  |
| Portion Eliminated (PE) | 0.71 | 0.44 | 0.98 |  |  |
| *Loneliness* |  |  |  |  |  |
| Controlled Direct Effect (CDE) | 1.09 | 0.64 | 1.55 |  |  |
| Pure Indirect Effect (PIE) | 1.15 | 0.95 | 1.35 |  |  |
| Reference Interaction (rINT) | 0.98 | 0.88 | 1.07 |  |  |
| Mediated Interaction (mINT) | 0.97 | 0.83 | 1.11 |  |  |
| Portion Eliminated (PE) | 1.09 | 0.87 | 1.32 |  |  |

**Table S26: Decomposition details for singles with children <16 years, compared to couples with children <16 years: Conservative Confounding**

|  | RR | 95% CI  (low) | 95% CI  (high) | Simulated proportion with psychiatric distress for singles with children | Simulated proportion with psychiatric distress for couples with children |
| --- | --- | --- | --- | --- | --- |
| *April 2020* |  |  |  |  |  |
| *Total Effect* | 1.26 | 0.95 | 1.57 | 0.54 | 0.43 |
| *Active Employment* |  |  |  |  |  |
| Controlled Direct Effect (CDE) | 1.26 | 0.86 | 1.65 |  |  |
| Pure Indirect Effect (PIE) | 1.00 | 0.99 | 1.02 |  |  |
| Reference Interaction (rINT) | 1.00 | 0.74 | 1.26 |  |  |
| Mediated Interaction (mINT) | 1.00 | 0.97 | 1.03 |  |  |
| Portion Eliminated (PE) | 1.00 | 0.75 | 1.25 |  |  |
| *Financial Difficulties* |  |  |  |  |  |
| Controlled Direct Effect (CDE) | 1.29 | 0.99 | 1.59 |  |  |
| Pure Indirect Effect (PIE) | 1.01 | 0.96 | 1.05 |  |  |
| Reference Interaction (rINT) | 0.97 | 0.91 | 1.03 |  |  |
| Mediated Interaction (mINT) | 1.00 | 0.97 | 1.02 |  |  |
| Portion Eliminated (PE) | 0.98 | 0.91 | 1.04 |  |  |
| *Childcare/Home-Schooling* |  |  |  |  |  |
| Controlled Direct Effect (CDE) | 1.52 | 0.97 | 2.06 |  |  |
| Pure Indirect Effect (PIE) | 0.99 | 0.97 | 1.02 |  |  |
| Reference Interaction (rINT) | 0.74 | 0.27 | 1.22 |  |  |
| Mediated Interaction (mINT) | 1.01 | 0.97 | 1.04 |  |  |
| Portion Eliminated (PE) | 0.74 | 0.28 | 1.21 |  |  |
| *Caring* |  |  |  |  |  |
| Controlled Direct Effect (CDE) | 1.09 | 0.74 | 1.44 |  |  |
| Pure Indirect Effect (PIE) | 1.01 | 0.99 | 1.03 |  |  |
| Reference Interaction (rINT) | 1.14 | 0.89 | 1.40 |  |  |
| Mediated Interaction (mINT) | 1.02 | 0.97 | 1.08 |  |  |
| Portion Eliminated (PE) | 1.17 | 0.88 | 1.47 |  |  |
| *Loneliness* |  |  |  |  |  |
| Controlled Direct Effect (CDE) | 1.21 | 0.91 | 1.51 |  |  |
| Pure Indirect Effect (PIE) | 1.09 | 0.99 | 1.20 |  |  |
| Reference Interaction (rINT) | 0.98 | 0.93 | 1.03 |  |  |
| Mediated Interaction (mINT) | 0.98 | 0.94 | 1.03 |  |  |
| Portion Eliminated (PE) | 1.06 | 0.94 | 1.17 |  |  |
| *January 2021* |  |  |  |  |  |
| *Total Effect* | 1.07 | 0.70 | 1.43 | 0.42 | 0.39 |
| *Active Employment* |  |  |  |  |  |
| Controlled Direct Effect (CDE) | 0.95 | 0.48 | 1.41 |  |  |
| Pure Indirect Effect (PIE) | 1.01 | 0.97 | 1.04 |  |  |
| Reference Interaction (rINT) | 1.16 | 0.77 | 1.55 |  |  |
| Mediated Interaction (mINT) | 0.98 | 0.91 | 1.05 |  |  |
| Portion Eliminated (PE) | 1.14 | 0.79 | 1.50 |  |  |
| *Financial Difficulties* |  |  |  |  |  |
| Controlled Direct Effect (CDE) | 1.14 | 0.77 | 1.52 |  |  |
| Pure Indirect Effect (PIE) | 1.04 | 0.97 | 1.11 |  |  |
| Reference Interaction (rINT) | 0.93 | 0.84 | 1.02 |  |  |
| Mediated Interaction (mINT) | 0.95 | 0.84 | 1.05 |  |  |
| Portion Eliminated (PE) | 0.92 | 0.79 | 1.04 |  |  |
| *Childcare/Home-Schooling* |  |  |  |  |  |
| Controlled Direct Effect (CDE) | 1.16 | 0.62 | 1.70 |  |  |
| Pure Indirect Effect (PIE) | 0.95 | 0.88 | 1.03 |  |  |
| Reference Interaction (rINT) | 0.98 | 0.47 | 1.48 |  |  |
| Mediated Interaction (mINT) | 1.00 | 0.89 | 1.12 |  |  |
| Portion Eliminated (PE) | 0.93 | 0.85 | 1.07 |  |  |
| *Caring* |  |  |  |  |  |
| Controlled Direct Effect (CDE) | 1.32 | 0.87 | 1.76 |  |  |
| Pure Indirect Effect (PIE) | 1.02 | 0.97 | 1.06 |  |  |
| Reference Interaction (rINT) | 0.81 | 0.63 | 0.98 |  |  |
| Mediated Interaction (mINT) | 0.92 | 0.78 | 1.05 |  |  |
| Portion Eliminated (PE) | 0.74 | 0.51 | 0.98 |  |  |
| *Loneliness* |  |  |  |  |  |
| Controlled Direct Effect (CDE) | 0.98 | 0.59 | 1.36 |  |  |
| Pure Indirect Effect (PIE) | 1.08 | 0.93 | 1.22 |  |  |
| Reference Interaction (rINT) | 0.98 | 0.87 | 1.09 |  |  |
| Mediated Interaction (mINT) | 0.99 | 0.91 | 1.07 |  |  |
| Portion Eliminated (PE) | 1.04 | 0.85 | 1.23 |  |  |

**Table S27: Decomposition details for singles with children <16 years, compared to singles with no children <16 years**

|  | RR | 95% CI  (low) | 95% CI  (high) | Simulated proportion with psychiatric distress for singles with children | Simulated proportion with psychiatric distress for singles with no children |
| --- | --- | --- | --- | --- | --- |
| *April 2020* |  |  |  |  |  |
| *Total Effect* | 1.24 | 0.91 | 1.58 | 0.54 | 0.44 |
| *Active Employment* |  |  |  |  |  |
| Controlled Direct Effect (CDE) | 1.23 | 0.82 | 1.63 |  |  |
| Pure Indirect Effect (PIE) | 1.00 | 0.98 | 1.02 |  |  |
| Reference Interaction (rINT) | 1.01 | 0.77 | 1.25 |  |  |
| Mediated Interaction (mINT) | 1.00 | 0.97 | 1.03 |  |  |
| Portion Eliminated (PE) | 1.01 | 0.76 | 1.25 |  |  |
| *Financial Difficulties* |  |  |  |  |  |
| Controlled Direct Effect (CDE) | 1.26 | 0.94 | 1.59 |  |  |
| Pure Indirect Effect (PIE) | 0.99 | 0.95 | 1.03 |  |  |
| Reference Interaction (rINT) | 0.98 | 0.91 | 1.06 |  |  |
| Mediated Interaction (mINT) | 1.00 | 0.98 | 1.03 |  |  |
| Portion Eliminated (PE) | 0.97 | 0.90 | 1.05 |  |  |
| *Childcare/Home-Schooling* |  |  |  |  |  |
| Controlled Direct Effect (CDE) | 1.25 | 0.75 | 1.76 |  |  |
| Pure Indirect Effect (PIE) | 1.15 | 0.86 | 1.44 |  |  |
| Reference Interaction (rINT) | 0.99 | 0.97 | 1.02 |  |  |
| Mediated Interaction (mINT) | 0.84 | 0.36 | 1.32 |  |  |
| Portion Eliminated (PE) | 0.98 | 0.58 | 1.38 |  |  |
| *Caring* |  |  |  |  |  |
| Controlled Direct Effect (CDE) | 1.01 | 0.66 | 1.37 |  |  |
| Pure Indirect Effect (PIE) | 1.00 | 0.97 | 1.03 |  |  |
| Reference Interaction (rINT) | 1.17 | 0.95 | 1.39 |  |  |
| Mediated Interaction (mINT) | 1.05 | 0.96 | 1.14 |  |  |
| Portion Eliminated (PE) | 1.22 | 0.94 | 1.51 |  |  |
| *Loneliness* |  |  |  |  |  |
| Controlled Direct Effect (CDE) | 1.24 | 0.91 | 1.58 |  |  |
| Pure Indirect Effect (PIE) | 0.99 | 0.91 | 1.07 |  |  |
| Reference Interaction (rINT) | 1.02 | 0.93 | 1.10 |  |  |
| Mediated Interaction (mINT) | 1.00 | 0.98 | 1.02 |  |  |
| Portion Eliminated (PE) | 1.01 | 0.89 | 1.12 |  |  |
| *January 2021* |  |  |  |  |  |
| *Total Effect* | 1.10 | 0.68 | 1.52 | 0.43 | 0.39 |
| *Active Employment* |  |  |  |  |  |
| Controlled Direct Effect (CDE) | 1.03 | 0.57 | 1.49 |  |  |
| Pure Indirect Effect (PIE) | 1.00 | 0.98 | 1.02 |  |  |
| Reference Interaction (rINT) | 1.09 | 0.74 | 1.44 |  |  |
| Mediated Interaction (mINT) | 0.99 | 0.93 | 1.05 |  |  |
| Portion Eliminated (PE) | 1.08 | 0.76 | 1.40 |  |  |
| *Financial Difficulties* |  |  |  |  |  |
| Controlled Direct Effect (CDE) | 1.16 | 0.75 | 1.58 |  |  |
| Pure Indirect Effect (PIE) | 1.03 | 0.96 | 1.10 |  |  |
| Reference Interaction (rINT) | 0.94 | 0.86 | 1.02 |  |  |
| Mediated Interaction (mINT) | 0.96 | 0.85 | 1.06 |  |  |
| Portion Eliminated (PE) | 0.93 | 0.81 | 1.05 |  |  |
| *Childcare/Home-Schooling* |  |  |  |  |  |
| Controlled Direct Effect (CDE) | 0.95 | 0.48 | 1.42 |  |  |
| Pure Indirect Effect (PIE) | 0.99 | 0.73 | 1.26 |  |  |
| Reference Interaction (rINT) | 1.01 | 0.98 | 1.03 |  |  |
| Mediated Interaction (mINT) | 1.17 | 0.76 | 1.57 |  |  |
| Portion Eliminated (PE) | 1.17 | 0.81 | 1.52 |  |  |
| *Caring* |  |  |  |  |  |
| Controlled Direct Effect (CDE) | 1.28 | 0.82 | 1.75 |  |  |
| Pure Indirect Effect (PIE) | 1.00 | 0.97 | 1.02 |  |  |
| Reference Interaction (rINT) | 0.83 | 0.64 | 1.02 |  |  |
| Mediated Interaction (mINT) | 0.97 | 0.88 | 1.07 |  |  |
| Portion Eliminated (PE) | 0.80 | 0.57 | 1.03 |  |  |
| *Loneliness* |  |  |  |  |  |
| Controlled Direct Effect (CDE) | 1.06 | 0.66 | 1.46 |  |  |
| Pure Indirect Effect (PIE) | 1.01 | 0.87 | 1.14 |  |  |
| Reference Interaction (rINT) | 0.99 | 0.84 | 1.15 |  |  |
| Mediated Interaction (mINT) | 1.00 | 0.94 | 1.05 |  |  |
| Portion Eliminated (PE) | 1.00 | 0.80 | 1.20 |  |  |

**Table S28: Decomposition details for singles with children <16 years, compared to singles with no children <16 years: Conservative Confounding**

|  | RR | 95% CI  (low) | 95% CI  (high) | Simulated proportion with psychiatric distress for singles with children | Simulated proportion with psychiatric distress for singles with no children |
| --- | --- | --- | --- | --- | --- |
| *April 2020* |  |  |  |  |  |
| *Total Effect* | 1.24 | 0.93 | 1.56 | 0.54 | 0.44 |
| *Active Employment* |  |  |  |  |  |
| Controlled Direct Effect (CDE) | 1.24 | 0.84 | 1.63 |  |  |
| Pure Indirect Effect (PIE) | 1.00 | 0.98 | 1.01 |  |  |
| Reference Interaction (rINT) | 1.01 | 0.78 | 1.24 |  |  |
| Mediated Interaction (mINT) | 1.00 | 0.97 | 1.03 |  |  |
| Portion Eliminated (PE) | 1.01 | 0.77 | 1.24 |  |  |
| *Financial Difficulties* |  |  |  |  |  |
| Controlled Direct Effect (CDE) | 1.28 | 0.97 | 1.59 |  |  |
| Pure Indirect Effect (PIE) | 0.98 | 0.94 | 1.02 |  |  |
| Reference Interaction (rINT) | 0.98 | 0.90 | 1.06 |  |  |
| Mediated Interaction (mINT) | 1.01 | 0.98 | 1.03 |  |  |
| Portion Eliminated (PE) | 0.97 | 0.89 | 1.04 |  |  |
| *Childcare/Home-Schooling* |  |  |  |  |  |
| Controlled Direct Effect (CDE) | 1.25 | 0.75 | 1.75 |  |  |
| Pure Indirect Effect (PIE) | 1.15 | 0.86 | 1.45 |  |  |
| Reference Interaction (rINT) | 0.99 | 0.97 | 1.02 |  |  |
| Mediated Interaction (mINT) | 0.84 | 0.36 | 1.33 |  |  |
| Portion Eliminated (PE) | 0.99 | 0.58 | 1.39 |  |  |
| *Caring* |  |  |  |  |  |
| Controlled Direct Effect (CDE) | 1.02 | 0.68 | 1.36 |  |  |
| Pure Indirect Effect (PIE) | 1.00 | 0.97 | 1.03 |  |  |
| Reference Interaction (rINT) | 1.17 | 0.95 | 1.39 |  |  |
| Mediated Interaction (mINT) | 1.06 | 0.96 | 1.15 |  |  |
| Portion Eliminated (PE) | 1.22 | 0.93 | 1.52 |  |  |
| *Loneliness* |  |  |  |  |  |
| Controlled Direct Effect (CDE) | 1.25 | 0.94 | 1.56 |  |  |
| Pure Indirect Effect (PIE) | 0.99 | 0.92 | 1.06 |  |  |
| Reference Interaction (rINT) | 1.02 | 0.93 | 1.10 |  |  |
| Mediated Interaction (mINT) | 1.00 | 0.98 | 1.02 |  |  |
| Portion Eliminated (PE) | 1.01 | 0.90 | 1.12 |  |  |
| *January 2021* |  |  |  |  |  |
| *Total Effect* | 1.19 | 0.78 | 1.61 | 0.42 | 0.35 |
| *Active Employment* |  |  |  |  |  |
| Controlled Direct Effect (CDE) | 1.12 | 0.63 | 1.61 |  |  |
| Pure Indirect Effect (PIE) | 1.00 | 0.97 | 1.03 |  |  |
| Reference Interaction (rINT) | 1.11 | 0.69 | 1.54 |  |  |
| Mediated Interaction (mINT) | 0.98 | 0.91 | 1.06 |  |  |
| Portion Eliminated (PE) | 1.10 | 0.72 | 1.48 |  |  |
| *Financial Difficulties* |  |  |  |  |  |
| Controlled Direct Effect (CDE) | 1.27 | 0.85 | 1.70 |  |  |
| Pure Indirect Effect (PIE) | 1.04 | 0.96 | 1.11 |  |  |
| Reference Interaction (rINT) | 0.93 | 0.83 | 1.02 |  |  |
| Mediated Interaction (mINT) | 0.95 | 0.83 | 1.06 |  |  |
| Portion Eliminated (PE) | 0.91 | 0.78 | 1.05 |  |  |
| *Childcare/Home-Schooling* |  |  |  |  |  |
| Controlled Direct Effect (CDE) | 1.03 | 0.51 | 1.54 |  |  |
| Pure Indirect Effect (PIE) | 0.99 | 0.69 | 1.30 |  |  |
| Reference Interaction (rINT) | 1.01 | 0.98 | 1.04 |  |  |
| Mediated Interaction (mINT) | 1.19 | 0.71 | 1.67 |  |  |
| Portion Eliminated (PE) | 1.19 | 0.78 | 1.59 |  |  |
| *Caring* |  |  |  |  |  |
| Controlled Direct Effect (CDE) | 1.41 | 0.91 | 1.90 |  |  |
| Pure Indirect Effect (PIE) | 1.00 | 0.97 | 1.02 |  |  |
| Reference Interaction (rINT) | 0.81 | 0.60 | 1.02 |  |  |
| Mediated Interaction (mINT) | 0.97 | 0.87 | 1.07 |  |  |
| Portion Eliminated (PE) | 0.77 | 0.53 | 1.02 |  |  |
| *Loneliness* |  |  |  |  |  |
| Controlled Direct Effect (CDE) | 1.15 | 0.72 | 1.59 |  |  |
| Pure Indirect Effect (PIE) | 1.02 | 0.89 | 1.15 |  |  |
| Reference Interaction (rINT) | 0.99 | 0.83 | 1.14 |  |  |
| Mediated Interaction (mINT) | 1.00 | 0.95 | 1.05 |  |  |
| Portion Eliminated (PE) | 1.00 | 0.79 | 1.21 |  |  |
